# Population size, HIV prevalence, and antiretroviral therapy coverage among key populations in sub-Saharan Africa: collation and synthesis of survey data 2010-2023

**DOI:** 10.1101/2022.07.27.22278071

**Authors:** Oliver Stevens, Keith Sabin, Rebecca Anderson, Sonia Arias Garcia, Kalai Willis, Amrita Rao, Anne F. McIntyre, Elizabeth Fearon, Emilie Grard, Alice Stuart-Brown, Frances Cowan, Louisa Degenhardt, James Stannah, Jinkou Zhao, Avi J. Hakim, Katherine Rucinski, Isabel Sathane, Makini Boothe, Lydia Atuhaire, Peter S. Nyasulu, Mathieu Maheu-Giroux, Lucy Platt, Brian Rice, Wolfgang Hladik, Stefan Baral, Mary Mahy, Jeffrey W. Imai-Eaton

## Abstract

**Background:** Key population HIV programmes in sub-Saharan Africa (SSA) require epidemiologic information to ensure equitable and universal access to effective services. We consolidated survey data among female sex workers (FSW), gay men and other men who have sex with men (MSM), people who inject drugs (PWID), and transgender people to estimate key population size, HIV prevalence, and antiretroviral therapy (ART) coverage for countries in mainland SSA.

**Methods:** Key population size estimates (KPSE), HIV prevalence, and ART coverage data from 39 SSA countries between 2010-2023 were collated from existing databases and verified against source documents. We used Bayesian mixed-effects spatial regression to model urban KPSE as a proportion of the gender/year/area-matched 15-49 years adult population. We modelled subnational key population HIV prevalence and ART coverage with age/gender/year/province-matched total population estimates as predictors.

**Findings:** We extracted 2065 key population size, 1183 HIV prevalence, and 259 ART coverage data points. Across national urban populations, a median of 1.65% of adult cisgender women were FSW (interquartile range [IQR]=1.35-1.91%), 0.89% of men were MSM (IQR=0.77-0.95%), 0.32% of men injected drugs (IQR=0.31-0.34%), and 0.10% of women were transgender (IQR=0.06-0.12%). HIV prevalence among key populations was, on average, 4 to 6 times higher than matched total population prevalence, and ART coverage was correlated with, but lower than, total population ART coverage with wide heterogeneity in relative ART coverage across studies. Across SSA, key populations were estimated as 1.2% (95% credible interval [CrI]: 0.9, 1.6) of the total population aged 15-49 years but 6.1% (95% CrI: 4.5, 8.2) of people living with HIV.

**Interpretation:** Key populations in SSA experience higher HIV prevalence and lower ART coverage, underscoring the need for focused prevention and treatment services. In 2024, limited data availability and heterogeneity constrain precise estimates for programming and monitoring trends. Strengthening key population surveys and routine data within national HIV strategic information systems would support more precise estimates.

**Funding:** UNAIDS, BMGF, NIH

**Research in Context:** *Evidence before this study:* Key populations, including female sex workers (FSW), gay men and other men who have sex with men (MSM), people who inject drugs (PWID), and transgender people, are at higher risk of HIV infection, including in sub-Saharan Africa (SSA). Delivering appropriate HIV prevention and treatment services for key populations and monitoring an equitable HIV response requires robust information on key population size, HIV prevalence, the treatment cascade, and new HIV infections. For this reason, key population surveys, including population size estimation and bio-behavioural surveys, are a standard component of comprehensive national HIV surveillance. Several complementary ongoing initiatives consolidate HIV data on key populations to support programme planning and implementation, global advocacy, and research. These include the Key Population Atlas and Global AIDS Monitoring (Joint United Nations Programme on HIV/AIDS [UNAIDS]), databases maintained by the US Centers for Disease Control and Prevention (CDC) and The Global Fund to Fight AIDS, Tuberculosis and Malaria (The Global Fund), and the Global.HIV initiative (Johns Hopkins University). These include similar data sources, but vary in scope, inclusion criteria, data elements recorded, and linkage to and validation against primary source reports. Incomplete recording of key methodological details limits appraisal and formal evidence synthesis, and therefore utility of data for strategic planning. Many other research studies have systematically reviewed, analysed, and extrapolated key population survey data in sub-Saharan Africa in single countries or across multiple countries. These studies have tended to focus on specific outcomes or population groups of interest, and primarily comprise an appraisal of peer-reviewed literature.

*Added value of this study:* We consolidated and deduplicated data collected between 2010-2023 from existing key population survey databases maintained by the UNAIDS Key Population Atlas, UNAIDS Global AIDS Monitoring, US CDC, and the Global Fund. We obtained published and grey literature surveillance reports from the Johns Hopkins University Global.HIV repository, additional web-based searches, and engagement with country HIV strategic information teams, and validated each observation of key population size, HIV prevalence, or ART coverage against primary surveillance reports. We used regression to characterise the relationship between key population and total population HIV indicators and extrapolated key population size estimates (KPSE), HIV prevalence, and ART coverage data to national-level estimates for all countries in mainland SSA. This exercise was the most comprehensive effort to date to consolidate key population HIV data in SSA. We analysed over 3000 observations from 126 KPSE, 217 HIV prevalence, and 62 ART coverage studies. We estimated that across urban populations aged 15-49 years in SSA countries, a median of 1.65% of cisgender women were FSW; 0.89% of men have sex with men; 0.32% of men injected drugs; and 0.10% of women were transgender. This translated to 3.7 million FSW, 1.9 million MSM, 770,000 PWID, and 230,000 transgender women (TGW) in SSA who require comprehensive HIV prevention or treatment services. FSW, MSM, PWID, and TGW together were estimated as 1.2% of the population aged 15-49 years, but comprised 6.1% of people living with HIV. ART coverage among members of key populations living with HIV increased with total population ART coverage, but was lower for all key populations. We identified large gaps in data availability. Of the four key populations and three indicators studied, only Mozambique had data for all twelve indicators. Data were particularly sparse for transgender populations and PWID.

*Implications of all the available evidence:* Key populations experience higher HIV prevalence and lower ART coverage across all settings in sub-Saharan Africa than the total population. Extrapolated national estimates provide a foundation for planning appropriate key population-focused services for HIV prevention and treatment in all settings, including those with no or limited data. However, large data availability gaps driven by discriminatory practices and punitive policies against key populations, inconsistency of existing data, and consequent wide uncertainty ranges around estimates limit the ability of existing data to guide granular programmatic planning and target setting for key population services and to monitor trends. More consistent surveillance implementation and improved routine surveillance through HIV prevention and treatment programmes for key populations would support monitoring equitable and equal programme access, as outlined in the Global AIDS Strategy 2021-2026 developed by UNAIDS, its co-sponsors, and other partners to end HIV/AIDS as a public health threat by 2030.

## Introduction

Key populations, including female sex workers (FSW), gay men and other men who have sex with men (MSM), people who inject drugs (PWID), and transgender people, experience higher risk of acquiring and transmitting HIV due to a combination of biological and sociostructural factors, including stigma and criminalisation.^1–3^ The Global AIDS Strategy 2021-2026 calls for “equitable and equal access” to HIV prevention and treatment to reduce HIV incidence and end HIV/AIDS as a public health threat by 2030.^4^ Delivering evidence-based HIV services for key populations and monitoring attainment of an equitable HIV response requires robust epidemiologic and service engagement indicators, including key population size, HIV prevalence and incidence, treatment and prevention cascades.

Key population survey data are sparse and less representative of their target population than common general population sampling approaches such as national household surveys or sentinel surveillance. Key population surveys often rely on respondent-driven and venue-based sampling methods^5,6^ which require strong assumptions to interpret as population-representative. Household survey sampling frames are inappropriate for reaching populations that are mobile and unlikely to disclose risk behaviour due to societal marginalisation and discrimination.^7–9^ Key population surveys are often infrequent and commonly restricted to, or disproportionately conducted in, urban areas.

Systematic reviews and meta-analyses of population size, HIV prevalence, or antiretroviral therapy (ART) coverage encompassing sub-Saharan Africa (SSA) have been conducted for FSW,^10,11^ MSM,^12,13^ PWID,^14,15^ and transgender women (TGW).^16,17^ Several independent initiatives have been undertaken to consolidate key population surveillance data.^18–21^ These efforts aim to monitor the state of the epidemic, evaluate programmes, and make recommendations on key population data reporting and quality thresholds.

We consolidated and harmonised key population size estimates (KPSE), HIV prevalence, and ART coverage data from existing databases and described data availability across countries and over time for each key population. We characterised the relationship between key population and total population HIV indicators and extrapolated KPSE, HIV prevalence, and ART coverage data to national-level estimates for mainland countries in SSA.

## Methods

### Data sources and extraction

We consolidated data from studies conducted 2010-2023 from four existing databases and two systematic reviews among FSW, MSM, PWID, and transgender people.^13,22^ Data among incarcerated people were not included as part of this review. The existing databases were the Joint United Nations Programme on HIV/AIDS (UNAIDS) Global AIDS Monitoring submissions,^23^ UNAIDS Key Population Atlas,^24^ the Global Fund to Fight AIDS, Tuberculosis and Malaria (the Global Fund) surveillance database, and a dataset maintained by the US Centers for Disease Control and Prevention (CDC) Division of Global HIV & TB Key Population Surveillance Team (Supplementary Table S1). For each observation, we extracted information about the study methodology, study location (country, subnational location), study population gender and age group, central estimate, sample size, standard error, and primary source or reference (e.g., survey report) (Supplementary Table S2).

Source documents were compiled from archives accompanying each database, the Johns Hopkins University Global.HIV document repository,^25^ Medline, and Google Scholar, following which we contacted UNAIDS Strategic Information advisors and other HIV programme contacts in each country to seek missing reports. Four authors (OS, RA, EG, AS-B) reviewed source documents to deduplicate observations recorded in multiple databases, validate against sources, and extract missing data elements. During source review, we added observations when additional relevant data were reported, or additional sources were ascertained that were not in the initial databases.

Observations were excluded if: key population definition, year, or study area were missing; the study area was non-specific (e.g., ‘urban areas’ or ‘5 provinces’); estimates were modelled or extrapolated; or data could not be confirmed by primary source review. There were no age group eligibility criteria for inclusion in analysis. Studies were not excluded or differentiated in our analysis according to study-specific population group definitions based on engaging in risk behaviours or time periods (e.g. sold sex in the last 6 months versus last 12 months; Supplementary Text S1). Transgender men (TGM) were not analysed or reported here due to insufficient data, but available observations are included in supplementary datasets (https://zenodo.org/records/10838438).

### Data processing

To compare population sizes across settings, KPSE reported as counts were converted to population proportions. Each KPSE was matched to the total population denominator^26–28^ by age, gender, year, and area, and assigned to one of seven method categories^29^ (Supplementary Text S2). Age information was missing from nearly all key population reports; missing age ranges were assigned as age 15-49 years. Unless gender was specified, PWID were assumed to be men as 90% of people who inject drugs in sub-Saharan Africa are men.^22^ Surveys among transgender people which did not report gender were assumed to be among transgender women because respondents often reported high prevalence of male partners and receptive anal intercourse, and surveys among TGM in SSA remain rare.^17^

Key population HIV prevalence and ART coverage observations were compared to total population HIV prevalence or ART coverage, respectively, matched by age, gender, year, and first administrative level (henceforth ‘province’). Provincial age/gender-specific HIV prevalence and ART coverage were extracted from UNAIDS Naomi subnational estimates for 2022 and projected backwards 2000-2021 parallel to national HIV prevalence and ART coverage trajectories among adults 15-49 years.^30^ Only serologically-determined HIV prevalence observations were included in analysis.

Key population ART coverage observations derived from populations with self-reported HIV-positive status or clinic-based recruitment were excluded. ART usage was determined through either self-report or laboratory methods via antiretroviral (ARV) metabolite biomarker or viral load (VL) testing. Viral load suppression (VLS) observations were standardised using a reporting threshold of 1000 copies/ml.^31^ In cases where surveys measured both ARV biomarker and VL, ARV biomarker was used. VLS observations were converted to estimates of ART coverage assuming a logit difference of -0.32, the average difference from studies that measured both viral load and metabolite-confirmed ARV status (Supplementary Text S3).

### Analysis

We used Bayesian mixed-effects linear regression to model logit-transformed KPSE proportions among the urban total population aged 15-49 years at provincial level. Separate regressions were estimated for each key population. The model included effects for study method, spatially-correlated random effects between neighbouring countries and provinces, and a study-level random effect allowing correlation among KPSE proportions from the same study. Study method was classified as: (1) non-mapping-based empirical methods that estimated total key population size, or (2) Priorities for Local AIDS Control Efforts (PLACE) or mapped estimates which counted venue- or hotspot-attending population size (see Supplementary Text S2 for details). Within these, method-specific random effects were included to permit empirical deviations from the central estimate. National KPSE counts were extrapolated using urban KPSE proportions, the proportion of the population living in urban areas,^32^ and the assumed rural-to-urban ratio of key population size. Qualitative data indicate that KPSE proportions are higher in urban areas than rural areas, but no empirical data were available to quantify the rural-to-urban ratio.^33–35^ To reflect wide uncertainty in the rural-to-urban ratio, we assumed that rural KPSE proportions were, on average, 40% lower than in urban areas, but with a wide range such that 80% of the time the true value will be between 20% and 60% lower.

For HIV prevalence, we modelled the relationship between logit-transformed key population HIV prevalence and logit total population HIV prevalence (age 15-49 years). FSW and PWID were modelled separately. Due to limited data, MSM and TGW were modelled together, but with a population type fixed effect reflecting empirical prevalence difference between TGW and MSM.^17^ We modelled survey HIV prevalence observations with a beta-binomial distribution to allow for overdispersion, accounting for additional variation between populations and study designs. The model included fixed effects for logit population prevalence interacted with region (Eastern and Southern Africa [ESA] or Western and Central Africa [WCA]) and province-, country-, and study-level random effects. Country- and province-level random effects were spatially correlated.

Primary analysis for key population ART coverage, was restricted to diagnostically-determined ART usage (either via ARV biomarker or VL testing). Logit-transformed key population ART coverage was modelled as a function of logit total population ART coverage in the same year, age group, and province. All key populations were modelled together with random slopes for each key population to enable sharing of information about the overall relationship for groups with very limited data. The observed number on ART was modelled with a beta-binomial distribution with fixed effects for logit population ART coverage, spatially correlated country- and province-level random effects, and study-level random effects.

We conducted sensitivity analyses in which self-reported ART coverage data were also included. For all three models (KPSE, prevalence, ART coverage) in sensitivity analysis we used ages 15-29 years as the matched total population denominator for MSM and TGW regression analyses, and 15-39 years for FSW, reflecting the younger median age of survey respondents.^36,–3736–38^

We combined national KPSE, HIV prevalence, and ART coverage estimates to calculate the number of KPLHIV and on ART. Ninety-five percent credible intervals were generated by combining 1000 posterior samples for each outcome. We calculated 1000 regional median KPSE differences from each of the posterior samples of national KPSE proportions, and calculated 95% credible intervals by combining the 1000 regional median differences. KPLHIV estimates were divided by estimates of the total PLHIV aged 15-49 years from 2022 national UNAIDS estimates to calculate the proportion of all PLHIV associated with each key population.^39^

Data were extracted, deduplicated, and validated in Microsoft Excel. Statistical analyses were conducted in R version 4.2.0 using the R-INLA package (v23.4.24). Further statistical methods details are in Supplementary Text S4. The GATHER reporting checklist^40^ is in Supplementary Table S17.

### Ethics statement

This study received ethical approval from the Imperial College Research Integrity and Governance Team (ICREC #6412027). Regulatory bodies providing ethical approvals or other ethics considerations for included studies are reported in Supplementary File S3.

## Results

4542 KPSE conducted between 2010 to 2023 were compiled (Figure 1). Following data cleaning, source document review, and area matching, 2065 observations were extracted from 126 studies (Table 1; Supplementary Figure S1). Data were most available for FSW (n=972, data from 35/39 countries; Supplementary Table S3), followed by MSM (n=647, 33/39 countries), PWID (n=329, 23/39 countries), TGW (n=117, 13/39 countries), and TGM (n=16, 2/39 countries). Sixty-nine percent (445/647) of urban MSM population proportion observations were below 1% of the male adult (15-49 years) population (Supplementary Figure S2).

**Figure 1:**
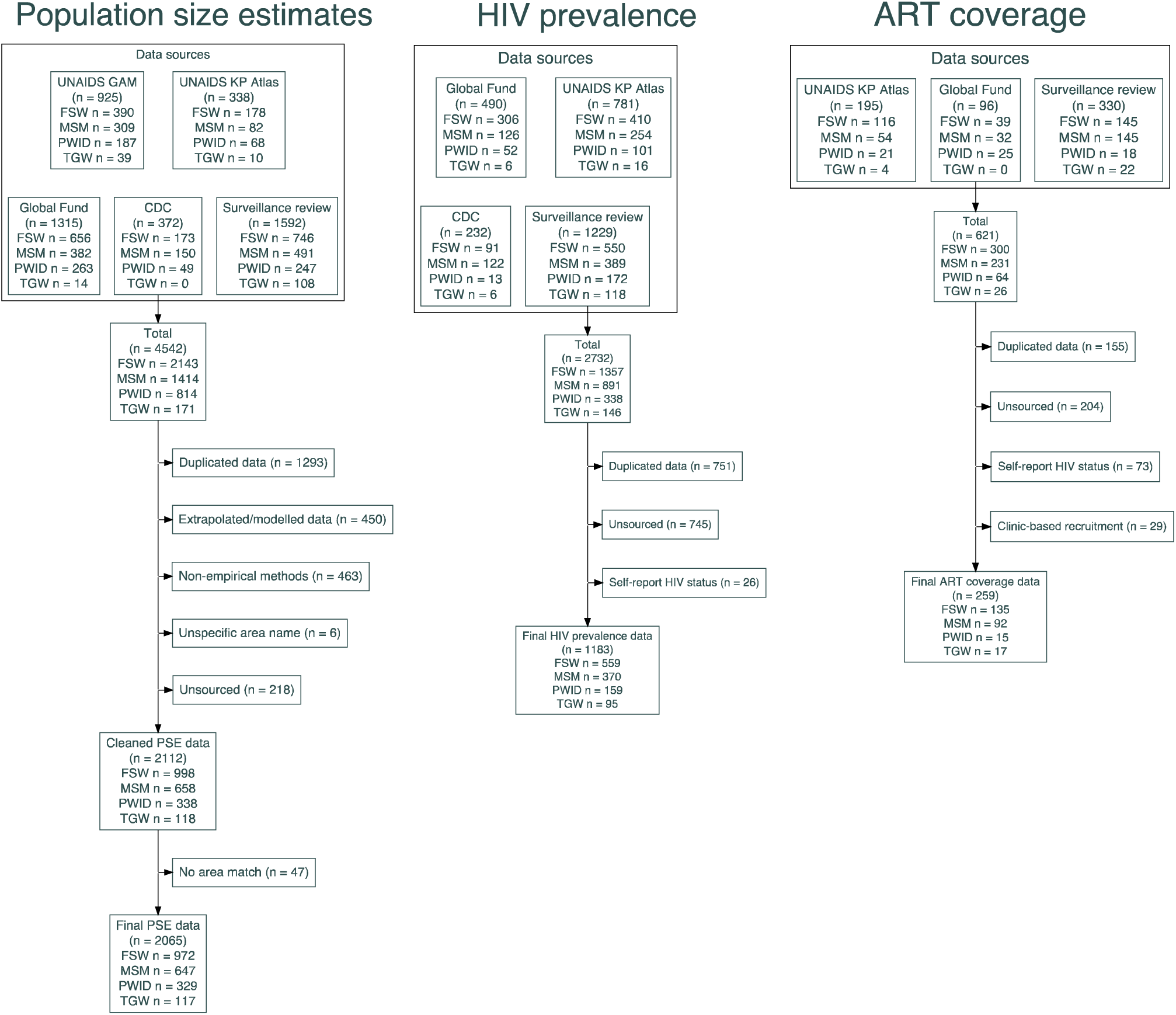
Flow diagrams describing identified data for key population size estimates, HIV prevalence data, and key population ART coverage. Each *n* represents number of population/method/location observations, with potentially observations for multiple subnational locations from the same study in same country. ART: Antiretroviral therapy coverage; FSW: Female sex worker; MSM: Men who have sex with men; PWID: People who inject drugs; TGW: Transgender women; KP: Key Population; GAM: Global AIDS Monitoring; CDC: US Centers for Disease Control and Prevention; PSE: Population size estimate.

**Table 1:** Availability of population size, HIV prevalence, and ART coverage data by key population and region during period 2010-2023.

| Region | Key population | Key Population Size Estimates |  | HIV prevalence |  | ART coverage |  |
| --- | --- | --- | --- | --- | --- | --- | --- |
|  |  | Data-points | Countries with data (n; %) | Data-points | Countries with data (n; %) | Data-points | Countries with data (n; %) |
| SSA<br>(39 countries) | FSW | 972 | 35 (90) | 559 | 36 (92) | 135 | 27 (69) |
|  | MSM | 647 | 33 (85) | 370 | 34 (87) | 92 | 24 (62) |
|  | PWID | 329 | 23 (59) | 159 | 21 (54) | 15 | 8 (21) |
|  | TGW | 117 | 13 (33) | 95 | 23 (59) | 17 | 10 (26) |
|  | TGM | 16 | 2 (5) | 5 | 5 (13) | 0 | 0 (0) |
| ESA<br>(18 countries) | FSW | 450 | 17 (94) | 236 | 18 (100) | 85 | 14 (78) |
|  | MSM | 291 | 15 (83) | 127 | 15 (83) | 45 | 12 (67) |
|  | PWID | 156 | 8 (44) | 52 | 9 (50) | 10 | 5 (28) |
|  | TGW | 65 | 7 (39) | 38 | 12 (67) | 13 | 7 (39) |
|  | TGM | 16 | 2 (11) | 2 | 2 (11) | 0 | 0 (0) |
| WCA<br>(21 countries) | FSW | 522 | 18 (86) | 323 | 18 (86) | 50 | 13 (62) |
|  | MSM | 356 | 18 (86) | 243 | 19 (90) | 47 | 12 (57) |
|  | PWID | 173 | 15 (71) | 107 | 12 (57) | 5 | 3 (14) |
|  | TGW | 52 | 6 (29) | 57 | 11 (52) | 4 | 3 (14) |
|  | TGM | 0 | 0 (0) | 3 | 3 (14) | 0 | 0 (0) |

SSA: Sub-Saharan Africa; ESA: East and Southern Africa; WCA: Western and Central Africa; FSW: female sex workers; MSM: men who have sex with men; PWID: people who inject drugs; TGW: transgender women; TGM: transgender men; ART: antiretroviral therapy

We identified 2732 key population HIV prevalence estimates, from which 1183 were extracted from 217 studies after processing. Denominators were reported for 98% (1154/1183) of observations. Most data were available for FSW (n=559, 36/39 countries), followed by MSM (n=370, 34/39 countries), PWID (n=159 21/39 countries), TGW (n=95, 23/39 countries), and TGM (n=5, 5/39 countries).

For ART coverage, 621 observations were identified. After processing, 259 observations were extracted from 62 studies. Denominators were available for 98% (254/259). Data were most available for FSW (n=135, 27/39 countries), followed by MSM (n=92, 24/39), TGW (n=17, 10/39 countries), and PWID (n=15, 8/39 countries). No data were available for TGM. Fifty-nine percent of observations were VLS [152/259], 9% ART metabolite testing [24/259], and 32% self-reported ART usage [83/259].

We estimated that, across countries, a median 1.65% of urban cisgender women aged 15-49 years were FSW (interquartile range [IQR] 1.35-1.91), 0.89% of men had sex with men (IQR 0.77-0.95), 0.32% of men injected drugs (IQR 0.31-0.34), and 0.10% of women were transgender (IQR 0.06-0.12; Figure 2). Incorporating the rural-to-urban ratio, median proportions of national total populations were 1.19% FSW, 0.66% MSM, 0.25% PWID, and 0.08% TGW. In sensitivity analysis using age 15-29 years as a denominator for MSM PSEs and 15-39 years for FSW PSEs, median urban proportions for MSM were 1.36% (IQR 1.15-1.42) and for FSW were 1.82% (IQR 1.60-2.13) (Supplementary Figure S3).

**Figure 2:**
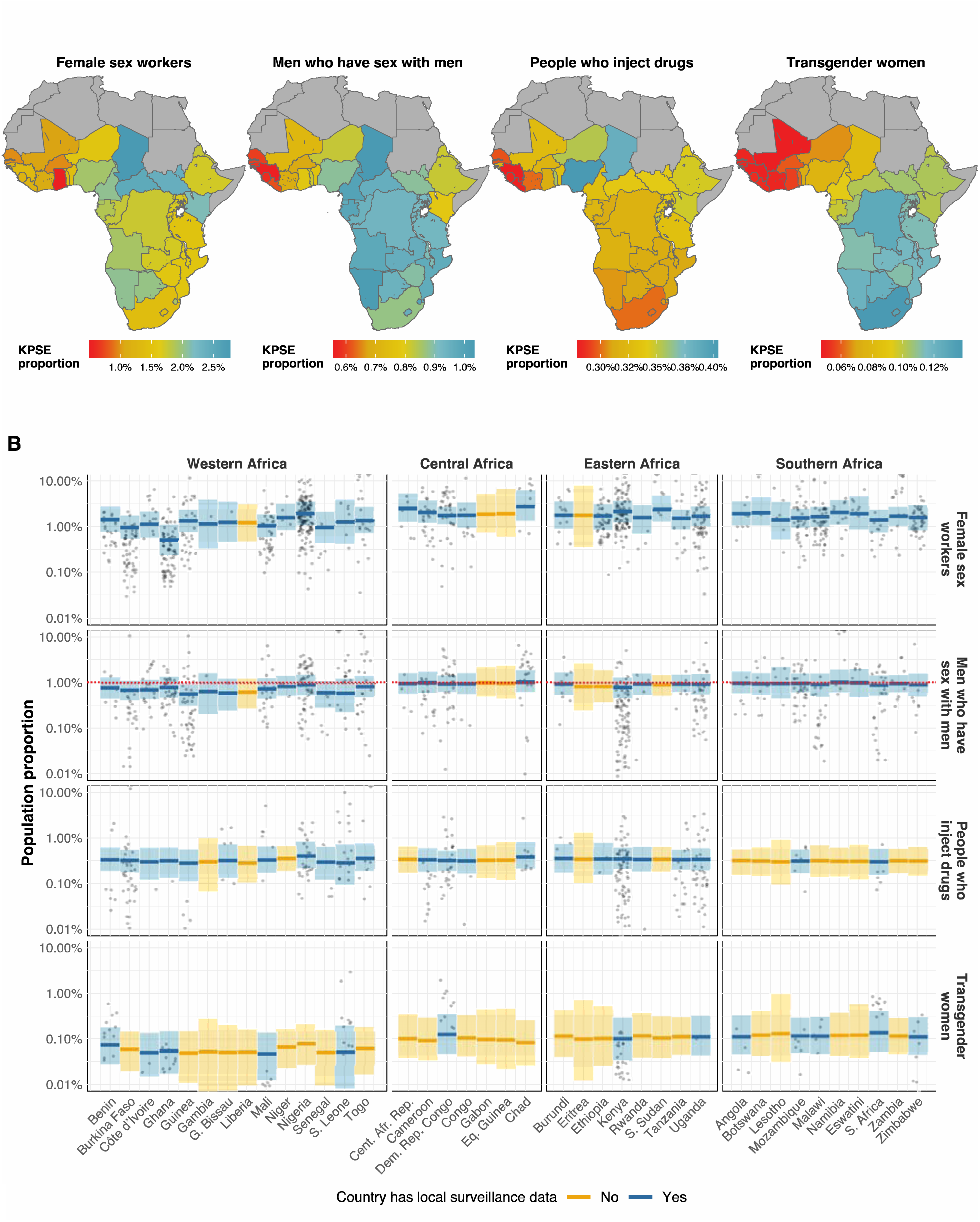
Model estimated urban key population size estimate (KPSE) proportions for female sex workers, men who have sex with men, people who inject drugs, and transgender women as a proportion of gender-matched 15-49 years adult total population. (A) Posterior median estimate for each country. Note: colour range is different for each key population chloropleth. (B) Posterior median estimate and 95% credible intervals for each country. Points represent observations of subnational KPSE proportions that used empirical methods. Countries that had local surveillance data are shown in blue, and countries informed only by spatial smoothing from neighbouring countries in yellow. Vertical axis shown on log-scale.

Urban KPSE proportions were higher in ESA than WCA among FSW (median difference +0.3%; 95% credible interval [CrI] -0.3, 1.0) and MSM (+0.2%; 95%CrI -0.2, 0.5) but uncertainty ranges contained zero. Proportions were similar between regions among PWID and TGW, but KPSE data were sparse overall. PSE proportions derived from PLACE and mapping studies were significantly lower than those derived from other methods among FSW (odds ratio 0.58; 95%CrI 0.40, 0.83), MSM (0.26; 95%CrI 0.17, 0.41), and PWID (0.31; 95%CrI 0.19, 0.53), and lower among TGW (0.27; 95%CrI 0.07, 1.11) but for TGW the 95% credible interval contained 1 (Supplementary Table S4; Supplementary Figure S4; Supplementary Tables S9-S12).

Observed HIV prevalence among key populations was consistently higher than area-matched total population HIV prevalence. Logit key population HIV prevalence in ESA was more strongly correlated with matched logit total population HIV prevalence than in WCA (Figure 3). In ESA, 15% population prevalence corresponded to 53% (95%CrI 45, 62) HIV prevalence among FSW, 21% (95%CrI 12, 31) among MSM, 28% (95%CrI 15, 45) among PWID, and 22% (95%CrI 14, 34) among TGW. In WCA, relative patterns were similar, but key population HIV prevalence varied less with total population HIV prevalence than in ESA. HIV prevalence differentials between key population and total population were larger when population prevalence was lower (Figure 3C). Supplementary Table S5-S7 report regression results, Supplementary Table S13 reports country-specific estimates, and Supplementary Figure S5 reports sensitivity analysis of imputed tested denominators.

**Figure 3:**
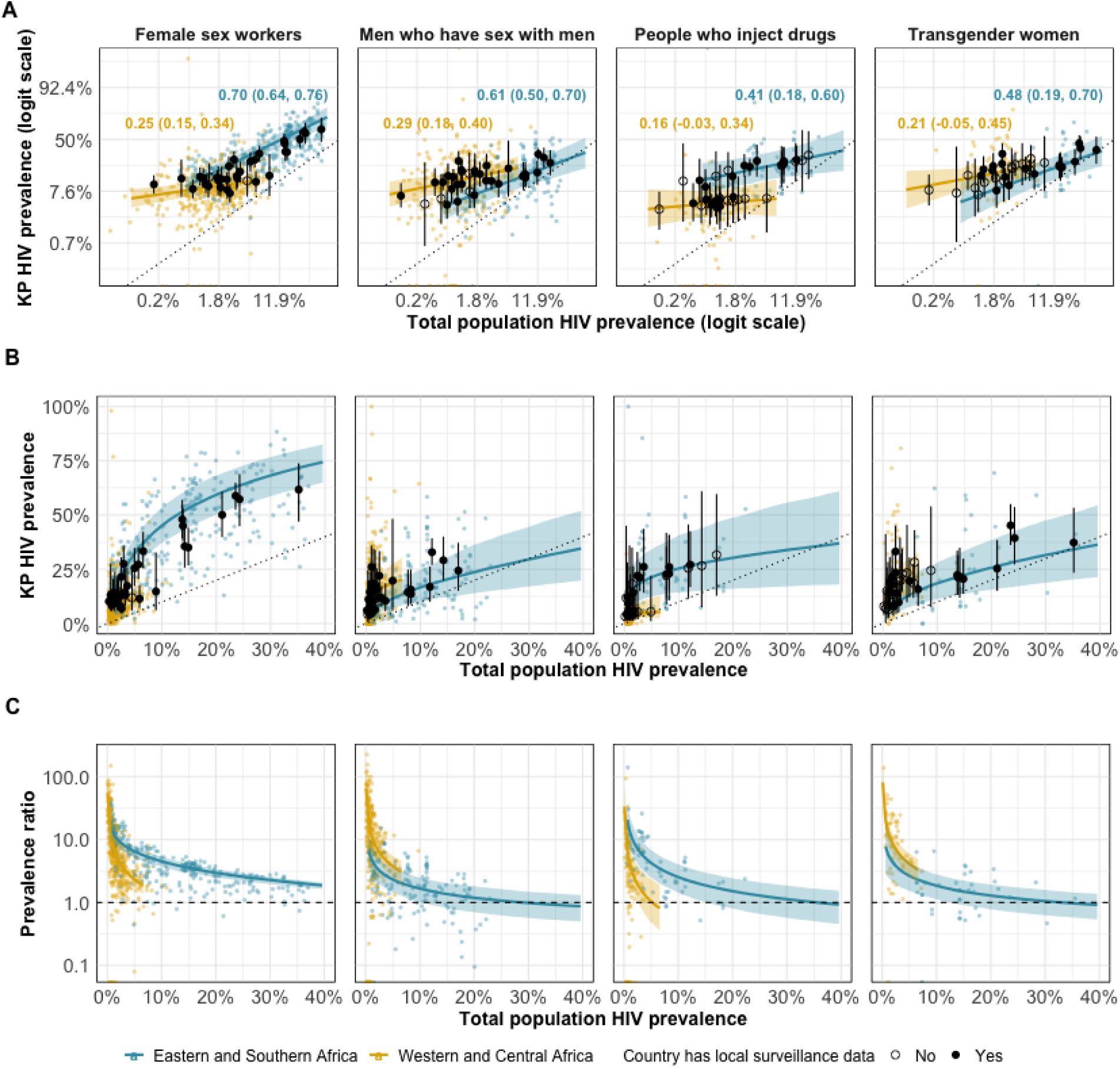
Key population and total population HIV prevalence on the logit scale (A), natural scale (B), and expressed as a ratio of key population and total population prevalence (C) for female sex workers, men who have sex with men, people who inject drugs, and transgender women. Coloured points indicate observed key population prevalence plotted against gender/year/province-matched total population prevalence. Logit-scale regional correlation coefficients and 95%CrI uncertainty intervals shown in coloured text in panel A. Coloured lines and shading represent regional estimate and 95% uncertainty results. Points represent country estimates [filled for countries with HIV prevalence data and empty for countries without HIV prevalence data] and 95% uncertainty ranges. Dotted line represents line of equality.

ART coverage among FSW, MSM, PWID, and TGW was correlated with gender-matched population ART coverage, but with wide heterogeneity in relative ART coverage across studies (Figure 4). At 80% population coverage, predicted ART coverage among FSW was 11% lower (95%CrI 19, 4%), 19% lower (30, 10%) among MSM, 25% lower (41, 11%) among PWID, 13% lower (43, 18%) among TGW. Supplementary Table S8 reports regression results and S14 reports country-specific estimates.

**Figure 4:**
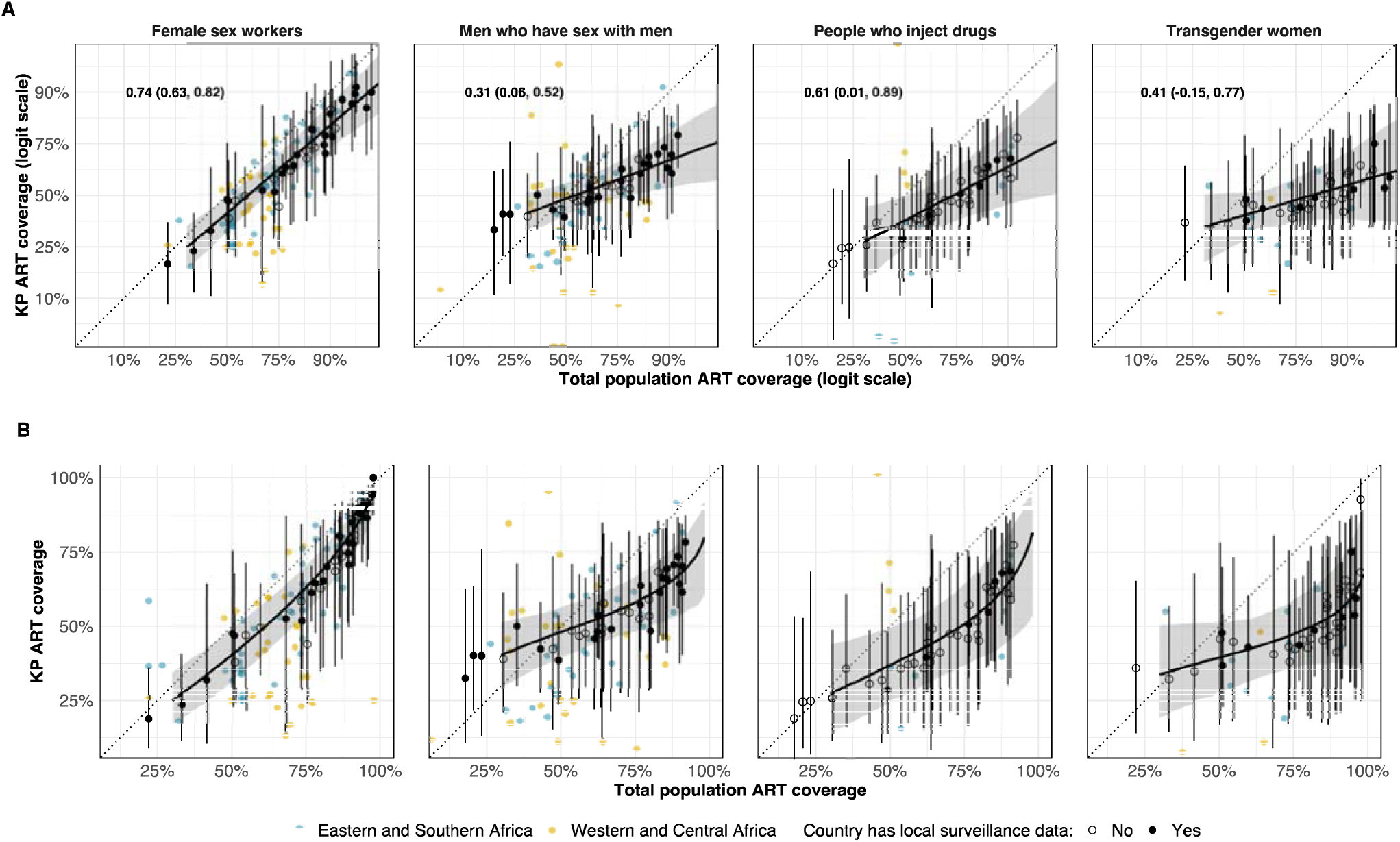
Estimated key population ART coverage as a function of total population ART coverage on the estimated logit scale (upper) and natural scale (lower). Coloured points indicate observed key population prevalence plotted against gender/year/province-matched total population ART coverage. Logit-scale correlation coefficients and 95%CrI uncertainty intervals shown in panel A. Black line and shading represent estimate for sub-Saharan Africa and 95% uncertainty results. Black points represent country estimates and 95% uncertainty ranges. Dotted line represents line of equality.

In sensitivity analysis including a fixed effect for ART measurement method, self-reported ART coverage was lower than lab-determined ART, but with wide 95% credible interval spanning no difference (odds ratio 0.73, 95%CrI 0.47, 1.15; Supplementary Figure S6, Table S8). Assuming MSM surveys represented MSM aged 15-29 years or FSW surveys represented FSW aged 15-39 years minimally affected on HIV prevalence or ART coverage estimates (Supplementary Table S16).

Across SSA, FSW, MSM, PWID, and TGW combined were 1.2% (95%CrI 0.9, 1.6) of the population aged 15-49 years (3.7m FSW, 1.9m MSM, 770,000 PWID, and 230,000 TGW), but 6.1% (95%CrI 4.5, 8.2) of PLHIV aged 15-49 years. In ESA this was 4.9% (95%CrI 3.4, 6.9) of PLHIV compared to 1.3% (95%CrI 0.9, 1.8) of the population and, in WCA, 11.7% (95%CrI 8.3, 16.5) of PLHIV compared to 1.2% (95%CrI 0.9, 1.7) of the population. FSW were 4.0% (95%CrI 2.7, 5.7) of PLHIV in SSA (750,000 PLHIV; 95%CrI 520,000, 1,100,000), MSM were 1.4% (95%CrI 0.9, 2.0) or 250,000 PLHIV (95%CrI 170,000, 380,000), PWID were 0.5% (95%CrI 0.3, 0.8) or 89,000 PLHIV (95%CrI 50,000, 150,000), and TGW were 0.2% (95%CrI 0.1, 0.5) or 44,000 PLHIV (95%CrI 20,000, 100,000) (Figure 5).

**Figure 5:**
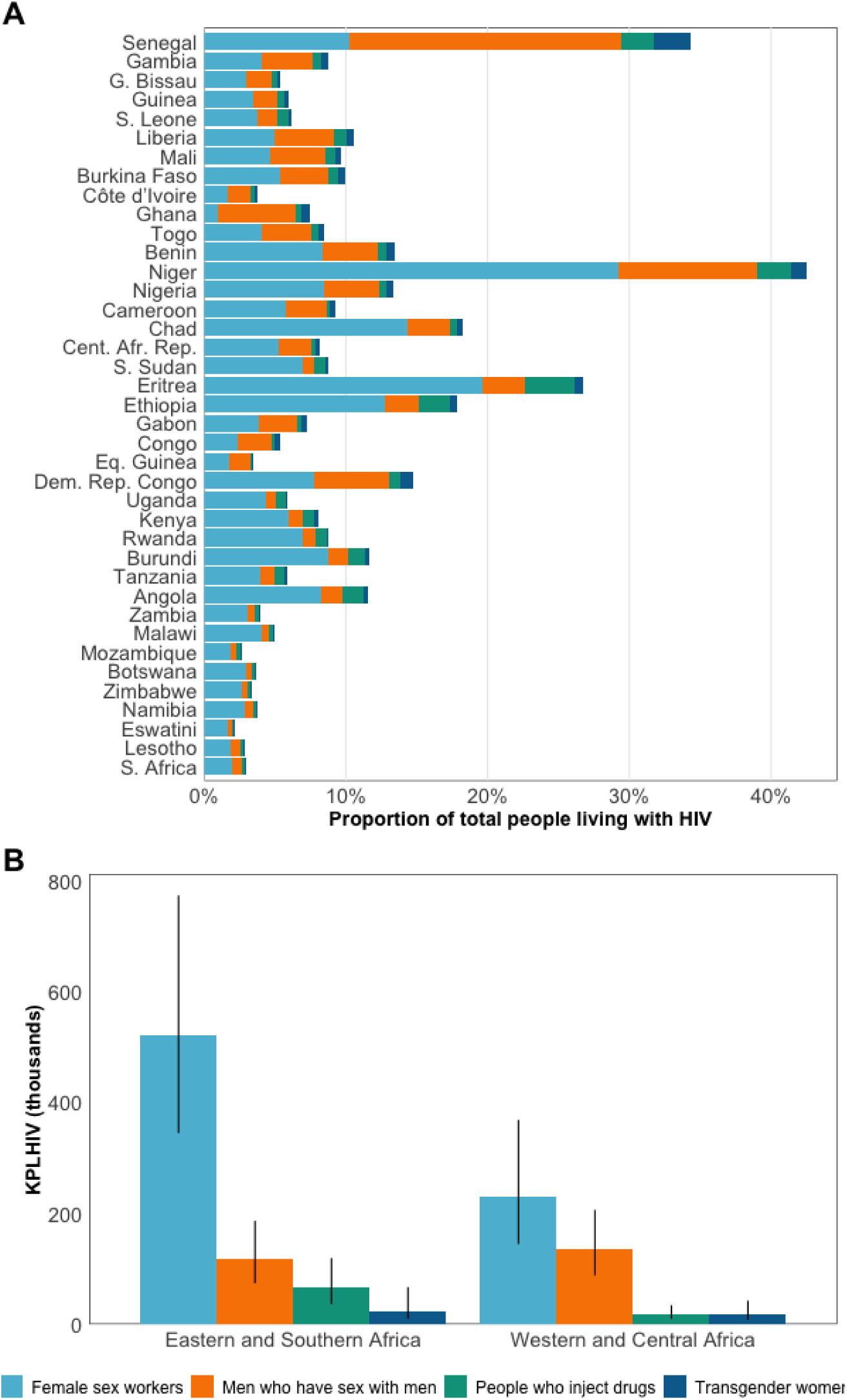
(A) HIV positive key population members as a proportion of all PLHIV aged 15-49 years. Countries are ordered geographically. (B) Estimated number of key populations living with HIV (KPLHIV) in Eastern and Southern Africa and Western and Central Africa.

## Discussion

Data from 273 key population surveys provided consistent evidence across sub-Saharan Africa of higher HIV prevalence and lower ART coverage among key populations. We estimated there are 3.7 million FSW, 1.9 million MSM, 770,000 PWID, and 230,000 TGW in SSA who require comprehensive HIV prevention or treatment services. HIV prevalence was estimated to be 21% among FSW, 14% among MSM, 11% among PWID, and 20% among TGW. These were 4.6, 5.9, 4.6, and 4.4 times higher than adult population prevalence, respectively, with larger relative differences when population prevalence was lower. Together, key populations constituted 1.2% of the adult population in SSA but 6.1% of PLHIV. ART coverage among key populations increased with total population coverage, but lagged behind at high population ART coverage levels. However, there was wide uncertainty about estimates aggregated within countries and across SSA.

This exercise highlighted major data gaps and challenges interpreting existing data. Across the twelve combinations of three indicators and four key populations analysed here, on average countries had data for seven combinations (Supplementary Table S3). Data gaps are larger when restricting to the highest quality and most recent data. These gaps impede planning for equitable and universal access to HIV services and will persist without firm commitments to collect key population survey data. Implementation of high-quality surveys will also require addressing entrenched discriminatory practices and punitive laws and policies against key populations^3,41–43^ In settings lacking or with limited data, our extrapolated estimates can be a foundation with which to guide future surveillance priorities, stimulate in-country data review and use, and estimate HIV epidemic indicators. Future efforts should especially focus on strengthening surveillance for MSM, PWID, and transgender people, and supporting use of HIV service delivery data for all key populations, while ensuring the safety and protection of marginalised individuals and their communities.^3^

Previous key population reviews and meta-analyses^11,44–46^ relied on systematic searches of peer-reviewed literature. Our initial sources were existing databases, primarily populated by surveillance teams in each country or implementing organisations. This resulted in including a larger grey literature body, such as survey reports, which are infrequently indexed by peer-reviewed databases.^47^ However, many observations from initial databases could not be validated in source documents and were excluded from analysis, which in some countries excluded the only reported data. Demand for total population HIV estimates in SSA has promoted high-quality comparable surveillance methods, documentation, and dissemination. Review of key population data by national HIV surveillance teams was introduced into the UNAIDS sub-Saharan African estimates process in 2022, providing context for existing surveys, identifying missing sources and adding new data. This, alongside ongoing key population data initiatives and concerted advocacy efforts that ensure public dissemination, will improve the foundation for HIV surveillance among key populations and promote parity for key populations within national HIV data and strategic information.

Systematically collated data on key population size, HIV prevalence, and ART coverage are critical inputs for mathematical modelling analyses that guide optimal HIV resource allocation. Such studies consistently demonstrate that fully meeting the HIV prevention and treatment needs of key populations would be impactful on reducing overall population incidence due to network effects of averting further onward transmission over time and to non-key population partners.^48,49^ Model-based counterfactual indicators that capture these impacts, such as the transmission preventable attributable fraction, are a better basis for intervention prioritisation than simple population proportions reported in survey data.^48,50,51^

Our estimates that 0.29% of men in SSA injected drugs was lower than recent estimates from Degenhardt *et al.* of 0.38%, though uncertainty ranges overlapped.^22^ This may reflect incorporating the urban-to-rural ratio in national PSE extrapolation in this analysis. We estimated that 1.2% of women aged 15-49 years sold sex, similar to the 1.1% estimated by Laga *et al.*^52^ Our finding of heterogeneous HIV prevalence ratios between MSM and the total population was consistent with Hessou *et al.*,^53^ particularly in WCA where HIV burden in neighbouring countries can vary considerably. Our estimate of 57% ART coverage among MSM in 2022 was lower than 73% from a meta-analysis by Stannah *et al.*^54^ of studies conducted up to 2022, with overlapping uncertainty. This may be due to exclusion of ART coverage using self-reported HIV status in this analysis.

Population size estimates measured in the same population at different times or with multiple methods were highly heterogenous, limiting data use for establishing meaningful programmatic targets or monitoring temporal trends. Very large uncertainty ranges around population size and PLHIV modelled estimates were consistent with findings of other statistically summarised and extrapolated PSE from multiple sources.^55–58^ Non-existence of rural KPSE studies to empirically inform rural-urban population proportions contributed to five-fold or greater relative uncertainty in extrapolated national estimates. Moreover, quality assessments of individual KPSE studies were not conducted as part of this analysis. Both systematically improving and evaluating study-specific implementation quality and methodological assumptions may reduce heterogeneity and increase the utility of KPSE data.^59–63^

Seventy-two percent of MSM KPSE observations and 38/39 modelled urban MSM estimates were below 1% of the male adult (15-49 years) population which UNAIDS and WHO recommended in 2020 as a minimum MSM population proportion.^64^ The minimum threshold was specified to ensure that MSM were not overlooked due to lack of data.

Variation in MSM proportions measured in PSE studies may reflect ability to engage in sexual behaviours or surveys, affected by legal and social environment and other factors, rather than differences in distribution of sexual preference. It is also uncertain whether MSM recruited in surveys, who are disproportionately young,^12,13^ are representative of the total MSM population. Specifying a minimum 1% threshold for MSM population could result in unrealistic MSM-focused service targets. The threshold should be reconsidered with the wider range of population size and stigma data now available, aligned with risk-based approaches to quantifying sex work and injecting drug use. Study-specific quality assessments will be important to producing accurate KPSE given stigma experienced by MSM in many settings.

Integrating key population programme data within key population estimates should be a priority to improve utility of surveillance data to guide programmatic response and monitor trends and subnational variation.^65^ Triangulating routine HIV testing and treatment data with biobehavioural survey data may help disentangle programmatic double counting, coverage of key population-specific services, and accounting for linkage to care in the estimation of ART coverage and viral load suppression.^66–68^ Future surveillance should consider moving beyond strictly defined key population definitions accurately to reflect heterogenous risks among individuals in overlapping risk environments including cisgender MSM and transgender women who sell sex, and FSW and MSM who inject drugs.^69–71^

Several limitations should be considered when interpretating these data and estimates. Firstly, surveys used a range of key population definitions and inclusion criteria which limited comparability, though there was sufficient overlap (Supplementary Text S2). Second, lack of explicit geographic survey sampling frames made it challenging to reflect the catchment population for a given survey in matched denominators, particularly given the mobility of key populations.^72^ This likely exacerbates the observed heterogeneity in KPSE proportions (Figure 2B), and is a major limitation for extrapolating KPSE proportions for programmatic planning. Going forward, specification of population catchments and careful data quality assessments to determine study inclusion and interpretation will improve future data use. Third, sample size or standard errors and age ranges were omitted from most population size data entries, gender was not specified for most observations among PWID, and a small number of observations were missing denominators for number tested. This limited ability to statistically weight, age-standardise, or gender-match data in model extrapolations, though sensitivity analyses indicate little impact on analysis conclusions. Fourth, data were insufficient and too heterogeneous to estimate time trends in population proportions, HIV prevalence, or ART coverage. Application of standardised recruitment methods, population definitions, and indicator reporting in consistent locations will improve monitoring of trends. Finally, this analysis did not include prisoners and other incarcerated people and future studies should look to expand data synthesis and analysis to this key population.

In conclusion, key populations across SSA experience disproportionate HIV burden and have lower antiretroviral therapy coverage. Consolidated key population data and synthesised estimates provide a basis for key population programming in all countries, including those with limited locally available data. Despite increasing focus on key populations in HIV/AIDS strategies in SSA, large data gaps remain and estimates are highly uncertain. New surveillance strategies, improved use of routine data, and more consistent surveillance implementation are required to furnish more precise estimates and trends for programme planning and support the monitoring of equitable and equal access to HIV prevention and treatment programmes outlined in the Global AIDS Strategy 2021-2026.

## Data Availability

All survey data produced in the present work are available at Zenodo link included in manuscript.
All remaining data are available upon reasonable request to the authors.

https://zenodo.org/records/10844137

## Authors’ contributions

OS, KS, MM and JWI-E conceptualised the study. KS, SAG, JZ, AMcI, KW, JS, LD curated key population survey databases. OS, RA, EG, AS-B reviewed primary source documents, deduplicated data, and extracted data. OS analysed the data and wrote the first draft of the manuscript. All authors contributed to interpretation of results and edited the manuscript for intellectual content. All authors read and approved the final version of the manuscript for submission.

## Disclaimer

The findings and conclusions in this report are those of the author(s) and do not necessarily represent the official position of the funding agencies.

## Data sharing

Data extracted and code to reproduce the analysis are available at https://zenodo.org/records/10838438.

## Acknowledgments

This research was supported by UNAIDS, Bill and Melinda Gates Foundation (INV-006733), National Institute of Allergy and Infectious Disease of the National Institutes of Health under award number R01AI136664, and the MRC Centre for Global Infectious Disease Analysis (reference MR/R015600/1), jointly funded by the UK Medical Research Council (MRC) and the UK Foreign, Commonwealth & Development Office (FCDO), under the MRC/FCDO Concordat agreement and is also part of the EDCTP2 programme supported by the European Union. MM-G’s research program is funded by a *Canada Research Chair* (Tier II) in *Population Health Modeling*.

## References

1. UNAIDS. The Gap Report.; 2014. Accessed April 1, 2022. https://www.unaids.org/en/resources/documents/2014/20140716_UNAIDS_gap_r eport

2. Garnett GP. Reductions in HIV incidence are likely to increase the importance of key population programmes for HIV control in sublJSaharan Africa. J Int AIDS Soc. 2021;24(S3):e25727. doi:10.1002/jia2.25727

3. World Health Organization. Consolidated Guidelines on HIV, Viral Hepatitis and STI Prevention, Diagnosis, Treatment and Care for Key Populations.; 2022. http://apps.who.int/iris/bitstream/10665/128048/1/9789241507431_eng.pdf?ua=1

4. United Nations. End Inequalities. End AIDS. Global AIDS Strategy 2021-2026 | UNAIDS. Published 2021. Accessed May 5, 2021. https://www.unaids.org/en/resources/documents/2021/2021-2026-global-AIDS-strategy

5. Baneshi MR, Rastegari A, Haghdoost AA. Review of Size Estimation Methods. In: Advances in Experimental Medicine and Biology. Vol 1333. Springer; 2021:1–15. doi:10.1007/978-3-030-75464-8_1

6. Abdul-Quader AS, Baughman AL, Hladik W. Estimating the size of key populations: Current status and future possibilities. Curr Opin HIV AIDS. Published online 2014. doi:10.1097/COH.0000000000000041

7. Jin H, Restar A, Beyrer C. Overview of the epidemiological conditions of HIV among key populations in Africa. J Int AIDS Soc. 2021;24(S3). doi:10.1002/jia2.25716

8. Hakim AJ, MacDonald V, Hladik W, et al. Gaps and opportunities: measuring the key population cascade through surveys and services to guide the HIV response. J Int AIDS Soc. 2018;21(Suppl Suppl 5):e25119. doi:10.1002/jia2.25119

9. Sabin K, Zhao J, Garcia Calleja JM, et al. Availability and quality of size estimations of female sex workers, men who have sex with men, people who inject drugs and transgender women in low- and middle-income countries. PLoS One. 2016;11(5). doi:10.1371/journal.pone.0155150

10. Atuhaire L, Adetokunboh O, Shumba C, Nyasulu PS. Effect of community-based interventions targeting female sex workers along the HIV care cascade in sub-Saharan Africa: a systematic review and meta-analysis. Syst Rev. Published online 2021. doi:10.1186/s13643-021-01688-4

11. Baral S, Beyrer C, Muessig K, et al. Burden of HIV among female sex workers in low-income and middle-income countries: A systematic review and meta-analysis. Lancet Infect Dis. Published online 2012. doi:10.1016/S1473-3099(12)70066-X

12. Stannah J, Dale E, Elmes J, et al. HIV testing and engagement with the HIV treatment cascade among men who have sex with men in Africa: a systematic review and meta-analysis. Lancet HIV. 2019;6(11):e769–e787. doi:10.1016/S2352-3018(19)30239-5

13. Stannah J, Soni N, Keng J, et al. Trends in HIV testing, the treatment cascade, and HIV incidence among men who have sex with men in Africa: A systematic review and meta-regression analysis. Lancet HIV. Published online 2023. doi:0.1016/S2352-3018(23)00111-X

14. Mathers BM, Degenhardt L, Phillips B, et al. Global epidemiology of injecting drug use and HIV among people who inject drugs: a systematic review. The Lancet. Published online 2008. doi:10.1016/S0140-6736(08)61311-2

15. Degenhardt L, Peacock A, Colledge S, et al. Global prevalence of injecting drug use and sociodemographic characteristics and prevalence of HIV, HBV, and HCV in people who inject drugs: a multistage systematic review. Lancet Glob Health. 2017;5(12):e1192–e1207. doi:10.1016/S2214-109X(17)30375-3

16. Kloek M, Bulstra CA, van Noord L, Al-Hassany L, Cowan FM, Hontelez JAC. HIV prevalence among men who have sex with men, transgender women and cisgender male sex workers in sub-Saharan Africa: a systematic review and meta-analysis. J Int AIDS Soc. 2022;25(11). doi:10.1002/JIA2.26022

17. Stevens O. HIV prevalence in transgender populations and cisgender men who have sex with men in sub-Saharan Africa 2010-2022: a meta-analysis. medRxiv. Published online 2023. doi:10.1101/2023.11.09.23298289

18. UNAIDS. Global AIDS Monitoring 2022. Published 2022. Accessed February 18, 2022. https://www.unaids.org/en/global-aids-monitoring

19. UNAIDS, WHO. Recommended Population Size Estimates for Men Who Have Sex with Men.; 2020.

20. Rao A, Schwartz S, Sabin K, et al. HIV-related data among key populations to inform evidence-based responses: protocol of a systematic review. Syst Rev. Published online 2018.

21. UNAIDS, FHI 360, WHO, CDC, PEPFAR. Biobehavioural Survey Guidelines For Populations At Risk For HIV.; 2017. Accessed February 17, 2022. http://apps.who.int/bookorders.

22. Degenhardt L, Webb P, Colledge-Frisby S, et al. Epidemiology of injecting drug use, prevalence of injecting-related harm, and exposure to behavioural and environmental risks among people who inject drugs: a systematic review. Lancet Glob Health. 2023;11(5):e659–e672. doi:10.1016/S2214-109X(23)00057-8

23. UNAIDS. Global AIDS Monitoring 2022. Published 2022. Accessed February 18, 2022. https://www.unaids.org/en/global-aids-monitoring

24. UNAIDS. Key Populations Atlas. Accessed November 27, 2021. https://kpatlas.unaids.org/dashboard

25. Rao A, Schwartz S, Sabin K, et al. HIV-related data among key populations to inform evidence-based responses: protocol of a systematic review. Syst Rev. Published online 2018.

26. UNAIDS. HIV sub-national estimates viewer. Published 2021. Accessed May 12, 2022. https://naomi-spectrum.unaids.org/

27. Eaton JW, Dwyer-Lindgren L, Gutreuter S, et al. Naomi: a new modelling tool for estimating HIV epidemic indicators at the district level in sub-Saharan Africa. J Int AIDS Soc. Published online 2021. doi:10.1002/jia2.25788

28. Global Rural-Urban Mapping Project (GRUMP). Urban boundaries. Accessed October 25, 2021. https://sedac.ciesin.columbia.edu/data/collection/grump-v1

29. Neal JJ, Prybylski D, Sanchez T, Hladik W. Population size estimation methods: Searching for the holy grail. JMIR Public Health Surveill. Published online 2020. doi:10.2196/25076

30. Eaton JW, Brown T, Puckett R, et al. The Estimation and Projection Package Age-Sex Model and the r-hybrid model: New tools for estimating HIV incidence trends in sub-Saharan Africa. AIDS. 2019;33(Suppl 3):S235–S244. doi:10.1097/QAD.0000000000002437

31. Johnson LF, Kariminia A, Trickey A, et al. Achieving consistency in measures of HIV-1 viral suppression across countries: derivation of an adjustment based on international antiretroviral treatment cohort data. J Int AIDS Soc. 2021;24 Suppl 5(Suppl 5). doi:10.1002/JIA2.25776

32. Nations U, of Economic D, Affairs S, Division P. World Urbanization Prospects The 2018 Revision.; 2018.

33. Maleke K, Makhakhe N, Peters RPH, et al. HIV risk and prevention among men who have sex with men in rural South Africa. African Journal of AIDS Research. 2017;16(1):31–38. doi:10.2989/16085906.2017.1292925

34. Wimark T. The City as a Single Gay Male Magnet? Gay and Lesbian Geographical Concentration in Sweden. 2014;752(September 2013):739–752.

35. Kobrak P, Ponce R, Zielony R. New Arrivals to New York CitylJ: Vulnerability to HIV among Urban Migrant Young Gay Men. Arch Sex Behav. Published online 2015:2041–2053. doi:10.1007/s10508-015-0494-4

36. Johnson LF, Mulongeni P, Marr A, Lane T. Age bias in survey sampling and implications for estimating HIV prevalence in men who have sex with men: Insights from mathematical modelling. Epidemiol Infect. 2018;146(8):1036–1042. doi:10.1017/S0950268818000961

37. Stannah J, Dale E, Elmes J, et al. HIV testing and engagement with the HIV treatment cascade among men who have sex with men in Africa: a systematic review and meta-analysis. Lancet HIV. 2019;6(11):e769–e787. doi:10.1016/S2352-3018(19)30239-5

38. Jones HS, Anderson RL, Cust H, et al. HIV incidence among women engaging in sex work in sub-Saharan Africa: a systematic review and meta-analysis. medRxiv. Published online January 1, 2023:2023.10.17.23297108. 10.1101/2023.10.17.23297108

39. UNAIDS. Global AIDS Update. Published 2021. Accessed May 12, 2022. https://www.unaids.org/en/resources/documents/2021/2021-global-aids-update

40. Stevens GA, Alkema L, Black RE, et al. Guidelines for Accurate and Transparent Health Estimates Reporting: the GATHER statement. The Lancet. 2016;388(10062):e19–e23. doi:10.1016/S0140-6736(16)30388-9

41. Lyons C, Bendaud V, Bourey C, et al. Global assessment of existing HIV and key population stigma indicators: A data mapping exercise to inform country-level stigma measurement. PLoS Med. 2022;19(2):e1003914. doi:10.1371/JOURNAL.PMED.1003914

42. UNAIDS. Prevailing against pandemics by putting people at the centre. Published online 2020.

43. HIV Policy Lab. HIV Policy Lab. Accessed October 25, 2023. https://www.hivpolicylab.org/

44. Hessou PHS, Glele-Ahanhanzo Y, Adekpedjou R, et al. Comparison of the prevalence rates of HIV infection between men who have sex with men (MSM) and men in the general population in sub-Saharan Africa: A systematic review and meta-analysis. BMC Public Health. 2019;19(1):1634. doi:10.1186/s12889-019-8000-x

45. Stutterheim SE, Van Dijk M, Wang H, Jonas KJ. The worldwide burden of HIV in transgender individuals: An updated systematic review and meta-analysis. PLoS One. 2021;16(12 December). doi:10.1371/journal.pone.0260063

46. Degenhardt L, Peacock A, Colledge S, et al. Global prevalence of injecting drug use and sociodemographic characteristics and prevalence of HIV, HBV, and HCV in people who inject drugs: a multistage systematic review. Lancet Glob Health. 2017;5(12):e1192–e1207. doi:10.1016/S2214-109X(17)30375-3

47. Stevens O. HIV prevalence in transgender populations and cisgender men who have sex with men in sub-Saharan Africa 2010-2021: a meta-analysis. Published online 2022.

48. Stone J, Mukandavire C, Boily M, et al. Estimating the contribution of key populations towards HIV transmission in South Africa. J Int AIDS Soc. 2021;24(1):e25650. doi:10.1002/jia2.25650

49. Mishra S, Pickles M, Blanchard JF, Moses S, Boily MC. Distinguishing sources of HIV transmission from the distribution of newly acquired HIV infections: Why is it important for HIV prevention planning? Sex Transm Infect. 2014;90(1):19–25. doi:10.1136/sextrans-2013-051250

50. Mishra S, Pickles M, Blanchard JF, Moses S, Shubber Z, Boily MC. Validation of the modes of transmission model as a tool to prioritize HIV prevention targets: A comparative modelling analysis. PLoS One. 2014;9(7). doi:10.1371/journal.pone.0101690

51. Shubber Z, Mishra S, Vesga JF, Boily MC. The HIV modes of transmission model: A systematic review of its findings and adherence to guidelines. J Int AIDS Soc. 2014;17(1). doi:10.7448/IAS.17.1.18928

52. Laga I, Niu X, Rucinski K, et al. Mapping the Population Size of Female Sex Worker in Countries Across Sub-Saharan Africa. PNAS. Published online September 19, 2023. doi:10.1073/pnas.2200633120

53. Hessou PHS, Glele-Ahanhanzo Y, Adekpedjou R, et al. Comparison of the prevalence rates of HIV infection between men who have sex with men (MSM) and men in the general population in sub-Saharan Africa: A systematic review and meta-analysis. BMC Public Health. 2019;19(1):1634. doi:10.1186/s12889-019-8000-x

54. Stannah J, Dale E, Elmes J, et al. HIV testing and engagement with the HIV treatment cascade among men who have sex with men in Africa: a systematic review and meta-analysis. Lancet HIV. Published online 2019. doi:10.1016/S2352-3018(19)30239-5

55. Edwards JK, Hileman S, Donastorg Y, et al. Estimating Sizes of Key Populations at the National Level: Considerations for Study Design and Analysis. Epidemiology. 2018;29(6):795–803. doi:10.1097/EDE.0000000000000906

56. Niu XM, Rao A, Chen D, et al. Using factor analyses to estimate the number of female sex workers across Malawi from multiple regional sources. Ann Epidemiol. 2021;55:34–40. doi:10.1016/j.annepidem.2020.12.001

57. Fearon E, Chabata ST, Magutshwa S, et al. Estimating the Population Size of Female Sex Workers in Zimbabwe: Comparison of Estimates Obtained Using Different Methods in Twenty Sites and Development of a National-Level Estimate. J Acquir Immune Defic Syndr. 2020;85(1):30–38. doi:10.1097/QAI.0000000000002393

58. Datta A, Lin W, Rao A, et al. Bayesian Estimation of MSM Population Size in Côte d’Ivoire. Statistics and Public Policy. 2019;6(1):1–13. doi:10.1080/2330443X.2018.1546634

59. Wesson PD, Mirzazadeh A, McFarland W. A Bayesian approach to synthesize estimates of the size of hidden populations: The anchored multiplier. Int J Epidemiol. 2018;47(5):1636–1644. doi:10.1093/ije/dyy132

60. Fellows I. Consensus Estimate Calculator. Accessed April 11, 2022. https://epiapps.com/shiny/app_direct/shinyproxy_combine_estimates/

61. USAID. Namibia Small Area Estimation: Final Report Small Area Estimation of Key Population Sizes in Namibia: Final Report.; 2021.

62. Datta A, Lin W, Rao A, et al. Bayesian Estimation of MSM Population Size in Côte d’Ivoire. Statistics and Public Policy. 2019;6(1):1–13. doi:10.1080/2330443X.2018.1546634

63. Rao A, Schwartz S, Viswasam N, et al. Evaluating the quality of HIV epidemiologic evidence for populations in the absence of a reliable sampling frame: a modified quality assessment tool. Ann Epidemiol. 2022;65:78–83. doi:10.1016/J.ANNEPIDEM.2021.07.009

64. UNAIDS, WHO. Recommended Population Size Estimates for Men Who Have Sex with Men.; 2020.

65. USAID. Namibia Small Area Estimation: Final Report Small Area Estimation of Key Population Sizes in Namibia: Final Report.; 2021.

66. Scheibe AP. Still left behind: Using programmatic data to assess harm reduction service coverage and HIV treatment cascades for people who inject drugs in five South African cities. In: IAS. ; 2019.

67. Scheibe A, Grasso M, Raymond HF, et al. Modelling the UNAIDS 90-90-90 treatment cascade for gay, bisexual and other men who have sex with men in South Africa: using the findings of a data triangulation process to map a way forward. AIDS Behav. 2018;22(3):853–859. doi:10.1007/s10461-017-1773-y

68. Hakim AJ, MacDonald V, Hladik W, et al. Gaps and opportunities: measuring the key population cascade through surveys and services to guide the HIV response. J Int AIDS Soc. 2018;21(Suppl Suppl 5):e25119. doi:10.1002/jia2.25119

69. Syvertsen JL, Agot K, Ohaga S, Bazzi AR. You can’t do this job when you are sober: Heroin use among female sex workers and the need for comprehensive drug treatment programming in Kenya. Drug Alcohol Depend. 2019;194:495–499. doi:10.1016/j.drugalcdep.2018.10.019

70. Poteat T, Ackerman B, Diouf D, et al. HIV prevalence and behavioral and psychosocial factors among transgender women and cisgender men who have sex with men in 8 African countries: A cross-sectional analysis. PLoS Med. Published online 2017. doi:10.1371/journal.pmed.1002422

71. Baral SD, Poteat T, Strömdahl S, Wirtz AL, Guadamuz TE, Beyrer C. Worldwide burden of HIV in transgender women: A systematic review and meta-analysis. Lancet Infect Dis. 2013;13(3):214–222. doi:10.1016/S1473-3099(12)70315-8

72. Davey C, Dirawo J, Mushati P, Magutshwa S, Hargreaves JR, Cowan FM. Mobility and sex work: why, where, when? A typology of female-sex-worker mobility in Zimbabwe. Soc Sci Med. 2019;220:322–330. doi:10.1016/j.socscimed.2018.11.027

