## Supplementary File 1 for "Population size, HIV prevalence, and antiretroviral therapy coverage among key populations in sub-Saharan Africa: collation and synthesis of survey data 2010-2023"

Supplementary Materials

Oliver Stevens, Keith Sabin, Sonia Arias Garcia, Rebecca Anderson, Kalai Willis, Amrita Rao, Anne McIntyre, Elizabeth Fearon, Emilie Grard, Alice Stuart-Brown, Frances Cowan, Louisa Degenhardt, James Stannah, Jinkou Zhao, Avi Hakim, Katherine Rucinski, Isabel Sathane, Makini Boothe, Lydia Atuhaire, Peter Nyasulu, Mathieu Maheu-Giroux, Lucy Platt, Brian Rice, Wolfgang Hladik, Stefan Baral, Mary Mahy, Jeffrey W. Imai-Eaton

*Correspondence to:*

Oliver Stevens

MRC Centre for Global Infectious Disease Analysis, School of Public Health

Imperial College London

St. Mary’s Hospital Campus

Norfolk Place

London W2 1PG,

United Kingdom

### Supplementary Text S1: Study population definitions and inclusion criteria based on risk behaviours and time periods

Inclusion criteria were extracted from 96 out of 247 studies. 79% of studies (76/96) reported an inclusion criterion based on engaging in risk behaviour within a specific recent time period and 88% (84/96) reported specific risk behaviours that defined inclusion in the population group.

Among FSW, 55% (17/31) required women to have sold sex in the last 6 months, followed by 29% (9/31) in the last 12 months, and 16% (5/31) in the last 1 month. Among MSM, 55% (17/31) required men to have had sex with men in the last 12 months, followed by 36% (11/31) in the last 6 months, and 10% (3/31) included men who had ever had sex with a man. Among PWID, 43% (6/14) specified injecting in the last 6 months, followed by 36% (5/14) in the last 12 months, 14% (2/14) in the last 3 months, and 7% (1/14) in the last month.

Among FSW, 68% (26/38) used a broad definition of sex work including exchanging sex for money, goods, or favours; 21% (8/38) required women to be commercial sex workers, or exchange sex for money only; and the remaining 4 surveys recruited women who self-identified as FSW or attended FSW clinics. Among MSM, all surveys required men to have had either anal sex (20%; 6/28) or oral or anal sex (80%; 24/28). Among PWID, 53% (8/15) surveys required injection of any drug, and a further 27% (4/15) specified injecting drug use of heroin, cocaine, or methamphetamine. The remaining surveys recruited people who used and injected drugs, from which this analysis extracted data on those who injected drugs.

### Supplementary Text S2: Key population size estimate method classifications

The five classifications for key population size estimates (KPSE) methods were:

- **2S-CRC:** two-source capture-recapture methods included object, service, and event multiplier methods.
- **3S-CRC:** three-source capture-recapture.
- **PLACE/mapping:** consisted of estimates using the *Priorities for Local AIDS Control Efforts* (PLACE) methodology and other programmatic/hotspot mapping derived estimates.
- **NSUM:** network scale-up methods.
- **SS-PSE:** successive sampling population size estimates, e.g. respondent-driven or snowball sampling.

Within a single survey, multiple size estimation methods were commonly conducted and combined into a final consensus KPSE; where possible, separate estimates for each method were extracted. For cases where only a final estimate of multiple methods was reported, two further categories were defined: **“Multiple methods – empirical”** or **“Multiple methods – mixture”**. The former contained estimates derived from multiple of the five methods above, while the latter were derived from both empirical and non-empirical methods (enumeration, wisdom of the crowds, key informant interviews, and the Delphi method). KPSEs derived by solely non-empirical methods were excluded from analyses.

### Supplementary Text S3: Viral load suppression observations

The definition of viral load suppression differed between studies. Most studies (n=25) used a threshold of ≤1000 copies/ml, one study each used thresholds of ≤500, ≤400, ≤398, and ≤200 copies/ml, and 5 studies used ≤50 copies/ml. Two studies (Nigeria 2020 BBS^1,2^ and Democratic Republic of Congo 2019 BBS) did not report a viral load suppression threshold and were assumed to be 1000 copies/ml. To standardise viral load suppression observations at 1000 copies/ml, we used the adjustment from Johnson et al. (2021)^3^*,* using the Weibull distribution with a shape parameter of 0.85.

Second, we converted viral load suppression estimates into estimates of ART coverage to facilitate comparison with total population ART coverage estimates. Five South African studies published both ART metabolite biomarker-confirmed ART usage and viral load suppression estimates. We meta-analysed these studies and estimated the average logit(ART) – logit(VLS) = -0.32. To convert viral load suppression observations into ART coverage, we used -0.32 as a regression offset.

1.	Nigeria Ministry of Health. *Integrated Biological & Behavioural Surveillance Survey (IBBSS) among Key Populations in Nigeria*.; 2020.

2.	Democratic Republic of Congo Ministry of Health. Enquete De Surveillance Bio-Comportementale Aupres Des Populations Cles A Risque De L’infection Par Le VIH Dans 14 Provinces, 2018-2019. Published online 2019:2018-2019.

3.	Johnson LF, Kariminia A, Trickey A, et al. Achieving consistency in measures of HIV-1 viral suppression across countries: derivation of an adjustment based on international antiretroviral treatment cohort data. *J Int AIDS Soc*. 2021;24 Suppl 5(Suppl 5). doi:10.1002/JIA2.25776

### Supplementary Text S4: Statistical model details

#### Key population size proportions

To facilitate comparison of population sizes across settings, KPSE reported as counts were converted to population proportions. Each KPSE was matched to the total population denominator by age, gender, year, and area. String matching was used to assign each survey area to geographic boundaries from the Global Rural/Urban Mapping Project (GRUMP) or UNAIDS Naomi subnational district populations. Where string matches were found for both GRUMP and Naomi subnational district populations, GRUMP populations were used.

We modelled urban key population sizes as a proportion of the total adult population aged 15-49 with a Bayesian spatial mixed-effects logistic regression model. Female sex workers were matched to women total population size denominator, men who have sex with men to men, people who inject drugs to both men, and transgender women to women. Spatial smoothing allowed population proportions to be correlated between geographically neighbouring provinces. Each key population was modelled with separate regression models. Logit population proportion was assumed to be normally distributed with mean $\mu_{i,x}$ (province $i\in1,2,...n_{provinces}$ and country $x\in1,2,\ldots, 39$) and standard deviation $\sigma$. The mean $\mu_{i,x}$ was modelled as a linear function including an intercept $\beta_{0}$, a fixed effect for study method, $X_{m}$ for $m\in\{\text{Empirical,}\text{ PLACE/Mapping}\text{\}}$using Empirical methods as the reference category, an independent and identically distributed (iid) random effect for study method, $\delta_{j}$ for $j\in\{\text{2S-CRC, 3S-CRC, network scaleup, SS-PSE, multiple methods (empirical), multiple methods (mixed)\},}$ an intrinsic conditional autoregressive (ICAR) spatial smoothing random effect at the provincial and national levels, $\theta_{i}$and $\eta_{x},$respectively, and a study iid random effect, $\epsilon_{s}$ for $s\in1,2,\ldots,n_{studies}$.

$$\mathrm{logit}\left( p_{i,x} \right)\sim N\left( \mu_{i,x},\sigma\right)$$

$$\mu_{i,x}=\beta_{0}+\beta_{1}X_{m}+{\delta_{j}+\theta}_{i}+\eta_{x}+\epsilon_{s}$$

$$\beta_{0},\beta_{1}\sim N\left( 0, 5 \right)$$

$$\theta_{i}\sim ICAR\left( \sigma_{\theta} \right)$$

$$\eta_{x}\sim ICAR\left( \sigma_{\eta} \right)$$

$$\delta_{j}\sim N(0, \sigma_{\delta})$$

$$\epsilon_{s}\sim N(0, \sigma_{\epsilon})$$

We then converted the PSE proportions into PSE counts:

$${PSE_{i, urban}= p}_{i} \times population_{i} \times urban{\%}_{i}$$

$$PSE_{i, rural}=\left( p_{i} \times R:U ratio \right) \times population_{i} \times(1-urban{\%}_{i})$$

The rural-to-urban ratio was assumed to be a $Beta(5,3)$ distribution, which had mean ratio of 0.6 and 80% of the mass between 0.4 and 0.8, allowing wide uncertainty to be propagated into the estimates of $PSE_{i, rural}$

#### Key population HIV prevalence

We modelled the relationship between key population HIV prevalence and sex-matched total population HIV prevalence (age 15-49 years) in the same admin-1 region separately for FSW and PWID, and together for MSM and TGW.

The number of key population members living with HIV, $X_{a,i,t,x,R}$, is assumed to follow a beta-binomial distribution with expected HIV prevalence, $p_{a,i,t,x,R}$ for a given age group $a$, province $i$, year $t,$ country $x$, and region $R\in\left\{ ESA, WCA \right\}.$ Logit transformed prevalence $logit(p_{a,i,t,x,R})$ is expressed a linear model with an intercept $\beta_{0}$, fixed effects for matched total population HIV prevalence $logit(\rho_{a,i,t,x,R})$; region $R \in ESA,WCA$; an interaction between matched total population HIV prevalence and region; an intrinsic conditional autoregressive (ICAR) spatial smoothing random effect at the provincial and national levels, $\theta_{i}$for $i\in1,2,...n_{provinces}$ and $\eta_{x} \text{for }x\in1,2\ldots39$ respectively, and a study *iid* random effect, $\epsilon_{s}$ for $s\in1,2,\ldots,n$.

$$X_{a,t,i,x,R}\sim BetaBinomial(n,p_{a,t,i,x,R})$$

$$logit\left( p_{a,t,i,x,R} \right)=\beta_{0}+\beta_{1}logit\left( \rho_{a,t,i,x,R} \right)+\beta_{2}R+\beta_{3}logit\left( \rho_{a,t,i,x,R} \right)\times R+\theta_{i}+\eta_{x}+\epsilon_{s}$$

$$\beta_{0},\beta_{1},\beta_{2},\beta_{3},\beta_{4}\sim N(0,5)$$

$$\theta_{i}\sim ICAR\left( \sigma_{\theta} \right)$$

$$\eta_{x}\sim ICAR\left( \sigma_{\eta} \right)$$

$$\epsilon_{s}\sim N(0,\sigma_{\epsilon})$$

HIV prevalence among TGW and MSM were modelled in a single regression. A fixed effect for KP $KP\in\{MSM, TGW\}$ and two interaction terms were added (matched total population HIV prevalence and key population; and region and key population) to permit KP-specific trends:

$$logit\left( p_{KP, a,t,i,x,R} \right)=\beta_{0}+\beta_{1}logit\left( \rho_{a,t,i,x,R} \right)+\beta_{2}R+\beta_{3}logit\left( \rho_{a,t,i,x,R} \right)\times R+\theta_{i}+\eta_{x}+\epsilon_{s}+\beta_{5}KP+\beta_{6}logit\left( \rho_{a,t,i,x,R} \right)\times KP+\beta_{7}\mathrm{KP}\times R$$

#### Key population ART coverage

We modelled key population ART coverage as a function of total population ART coverage, analogously to HIV prevalence. All key populations were modelled together.

The observed number on ART, $A_{KP, a,t,x,i}$, was assumed to have a beta-binomial distribution with the probability of being on treatment, $p_{KP, a,t,x,i}$ for a given key population $KP,$age group $a$, year $t$, country $x$, and province $i$. ${logit(p}_{KP, a,t,x,i})$ was modelled as a linear equation with intercept $\beta_{0}$, fixed effect for matched total population ART coverage, random intercept for key population $KP\in\{FSW, MSM, PWID, TGW\}$, random slopes matched total population ART coverage by key population, an intrinsic conditional autoregressive (ICAR) spatial smoothing random effect at the provincial and national levels, $\theta_{i}$for $i\in1,2,\ldots, n_{provinces}$ and $\eta_{x} \text{for }x\in1,2,\ldots,39$ respectively, and a study *iid* random effect, $\epsilon_{s}$ for $s\in1,2, \ldots, n$. Insufficient data were available to estimate region-specific trends.

$$A_{KP, a,t,x,i}\sim BetaBinomial(n,p_{a,t,x,i})$$

$$logit\left( p_{KP, a,t,x,i} \right)=\beta_{0}+\beta_{1}logit\left( \alpha_{a,t,x,i} \right)+u_{0KP}+u_{1KP}logit\left( \alpha_{a,t,x,i} \right)+{\theta_{i}+\eta_{x}+\epsilon}_{s}$$

$$\beta_{0},\beta_{1}\sim N(0,5)$$

$$u_{0KP}\sim N\left( 0, {\sigma_{u}}_{0} \right)$$

$$u_{1}\sim N(0, {\sigma_{u}}_{1})$$

$$\theta_{i}\sim ICAR\left( \sigma_{\theta} \right)$$

$$\eta_{x}\sim ICAR\left( \sigma_{\eta} \right)$$

$$\epsilon_{s}\sim N(0,\sigma_{\epsilon})$$

In sensitivity analysis, we added a fixed effect for study method, $M$ where $M=0$ for laboratory confirmed ART and $M =1$ for self-reported ART usage.

National gender-matched ART coverage in 2022 from Spectrum files were used to estimate national key population ART coverages (Supplementary Table S11). Spectrum estimates gender-specific ART coverage by dividing gender-specific ART programme counts over the gender-specific estimate of the number of people living with HIV. Misspecification of the sex ratio of new HIV infections can lead to an under-enumeration of women living with HIV and an over-enumeration of men living with HIV. This leads to an overestimate of ART coverage among women, and an underestimate of ART coverage among men. We assessed this to have occurred in Benin, Burkina Faso, Malawi, Senegal, and Sierra Leone which had implausibly large differences between female and male ART coverage, defined as $logit\left( \text{ART }\text{coverage}_{female} \right)-logit\left( \text{ART }\text{coverage}_{male} \right)>2$. We adjusted the logit difference of gender-specific ART coverages in these countries to be 0.72, the median logit difference across the remaining 34 countries in SSA.

**Supplementary Figures and Tables**

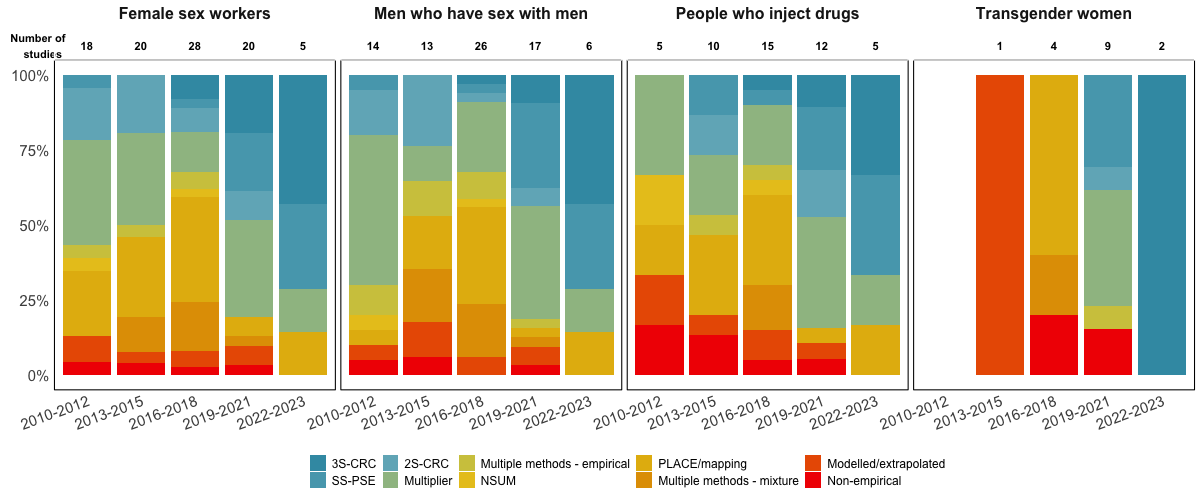

Supplementary Figure S1: Distribution of size estimation methods implemented over time by key population. The number of size estimation studies is shown above the bars. Bars are colour coded by method empiricism from highest (3S-CRC) to lowest (non-empirical). Population size estimation studies using empirical methods have become more common since 2010. Between 2019-2021, 20 FSW, 18 MSM, 11 PWID, and 7 TGW PSE studies were conducted, increasing from 15, 12, 4, and 0, respectively, in 2010-12.

3S-CRC: Three-source capture-recapture; 2S-CRC: Two-source capture-recapture; SS-PSE: Successive sampling population size estimate; NSUM: network scale-up method; PLACE: Priorities for Local AIDS Control Efforts

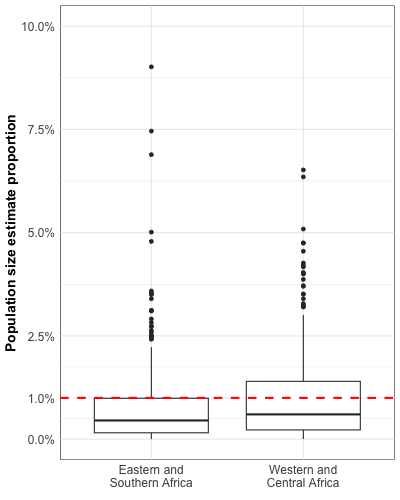

Supplementary Figure S2: Empirical observations of men who have sex with men (MSM) population size estimate proportions. Observations stratified by Eastern and Southern Africa (n = 360) and Western and Central Africa (n = 294). Boxplots represent median and interquartile range of observations. Red dashed line is the UNAIDS/WHO recommended 1% threshold for MSM population proportions. UNAIDS: Joint United Nations Programme on HIV/AIDS; WHO: World Health Organization

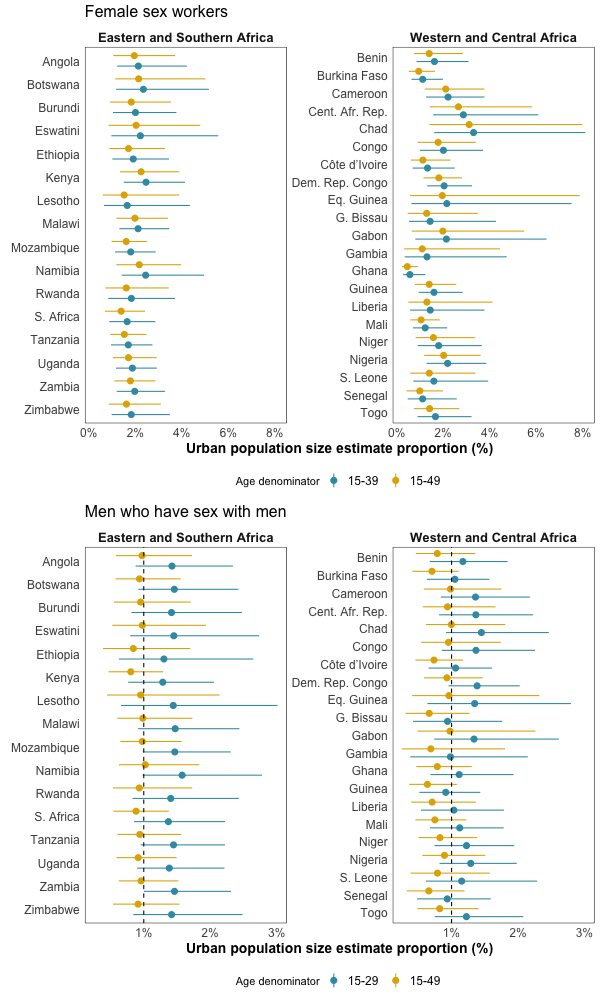

Supplementary Figure S3: Age group sensitivity analysis for FSW and MSM population proportion. Key population surveys may recruit individuals younger than the 15-49 year old denominator as assumed in primary analysis (yellow; main text Figure 3B). This sensitivity analysis estimates urban PSE proportions for MSM using all men aged 15-29 as the matched total population denominator (blue), and for FSW using all women aged 15-39. This increases the PSE proportions as the denominator has decreased. The dotted line on the MSM plot represents the UNAIDS/WHO recommended minimum population size proportion of 1% of total population men. MSM: Men who have sex with men

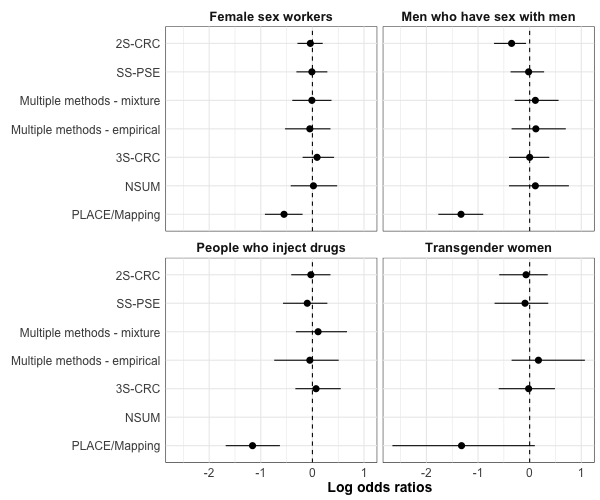

Supplementary Figure S4: Population size method effects. Log odds ratios for population size estimates by methods. Two fixed effect categories were estimated: Empirical and PLACE/Mapping. Random effects were estimated for empirical methods (Two and three source CRC (2S- and 3S-CRC), network scale-up (NSUM), successive sampling population size estimation (SS-PSE), and average estimates from multiple empirical methods and estimates derived from a mixture of empirical and non-empirical methods (Multiple methods – mixed)). See Supplementary Table S4 for a tabular representation of population size estimate method effects. Each key population was estimated in separate regression models, but results presented together to enable comparison of estimates. PLACE: Priorities for Local AIDS Control Efforts; FSW: female sex workers; MSM: men who have sex with men; PWID: people who inject drugs; TGW: transgender women

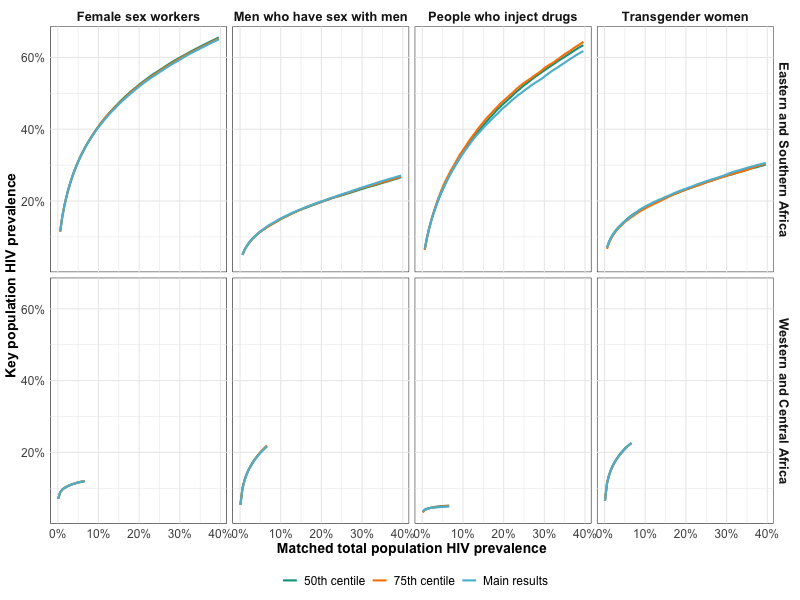

Supplementary Figure S5: Sensitivity analysis for imputed tested denominators of HIV prevalence observations. The main results present estimates with observations missing tested denominators imputed using the 25^th^ centile of key population-matched known denominators. This sensitivity analysis shows two further imputations, using the median (50^th^ centile) of known denominators, and the 75^th^ centile.

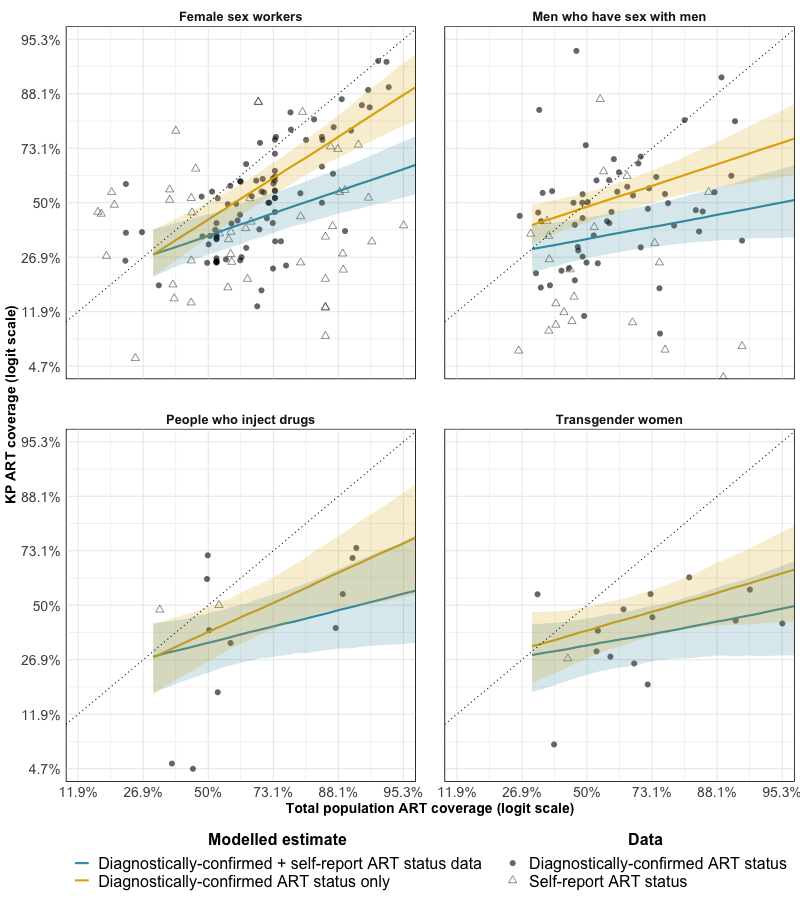

Supplementary Figure S6: ART coverage model sensitivity analysis. Yellow estimates are as in primary analysis, omitting a fixed effect for study method (self-report or diagnostically confirmed ART status). Blue estimates include self-reported ART status data with a diagnostically-confirmed HIV status denominator and a fixed effect for study method.

### Supplementary Table S1: Key population databases

| Database | Access | Years extracted | Description | KP indicators captured | Data entry |
| --- | --- | --- | --- | --- | --- |
| UNAIDS Global AIDS Monitoring (GAM) | <https://www.unaids.org/en/global-aids-monitoring> | 2010-2022 | Global monitoring tool to track progress towards key targets and assess state of HIV epidemic | PSE, HIV prevalence, ART coverage/viral load suppression. Programme HIV prevention data on condoms, opioid substitution therapy, and needle and syringe programmes.  Raw data or modelled estimates | National government annual data submission |
| UNAIDS KP Atlas | <https://kpatlas.unaids.org> | 2010-2022 | Consolidated database maintained by UNAIDS, supported by Global Fund, WHO, US CDC | PSE, HIV prevalence, ART coverage, HIV testing, HIV prevention data, laws and stigma indices  Raw data or modelled estimates | UNAIDS-maintained. Aligned with GAM since 2017 |
| Global Fund | Private (Contact Jinkou Zhao) | 2010-2017 | Global Fund surveillance database | PSE, HIV prevalence, ART coverage, HIV prevention  Raw data or modelled estimates | National governments through grant reporting. In recent years, aligned with KP Atlas and GAM |
| CDC | Private  (Contact Avi Hakim) | 2010-2022 | CDC surveillance spreadsheet | Majority PSE, small amount of HIV prevalence and ART coverage data  Raw data or modelled estimates | CDC maintained |
| Global.HIV repository | <https://www.ncbi.nlm.nih.gov/pmc/articles/PMC6278072/> | 2010-2017 | Systematic review database | Wide range of epidemiological, human rights, and healthcare indicators  Raw data or modelled estimates | Maintained by Johns Hopkins University |
| Degenhardt et al. PWID systematic review | <https://pubmed.ncbi.nlm.nih.gov/29074409/> | 2010-2017 | Systematic review database | PSE, HIV prevalence, ART coverage for PWID  Raw data only | Maintained by University of New South Wales |
| Stannah et al. MSM systematic review | <https://pubmed.ncbi.nlm.nih.gov/31601542/> | 2010-2018 | Systematic review database | HIV prevalence, ART coverage for MSM  Raw data only | Maintained by McGill University |
| Stannah et al. MSM systematic review | <https://www.medrxiv.org/content/10.1101/2022.11.14.22282329v1> | 2010-2022 | Systematic review database | HIV prevalence, ART coverage for MSM  Raw data only | Maintained by McGill University |

### Supplementary Table S2: Extracted data elements

| Data element | Population size estimate (PSE) data | HIV prevalence data | ART coverage data |
| --- | --- | --- | --- |
| Country | Country name | Country name | Country name |
| Key population | FSW; MSM; PWID; TGW; TGM | FSW; MSM; PWID; TGW; TGM | FSW; MSM; PWID; TGW; TGM |
| Year | Year of study | Year of study | Year of study |
| Method | Methods employed to estimate population size.  One of:   - Two source capture recapture (2S-CRC) - Three source capture recapture (3S-CRC) - Event multiplier - NSUM - Object multiplier - PLACE/mapping - Service multiplier - SS-PSE - Delphi - Wisdom of the crowds - Enumeration - Key informant interviews - Literature estimates   When several methods were used to create a median or consensus estimates, the individual method estimates were recorded, and the median estimate was not. In cases where only the median was reported two further categories are defined:   - Multiple methods - empirical:  All methods used to create the median estimate were from the eight methods listed above. - Multiple methods - mixture:  Methods used to create the median estimate were a mixture of one or more of the eight methods above, plus a non-empirical method (e.g. wisdom of the crowds, enumeration, literature review). | Methods employed to assess HIV prevalence.  One of:   - Laboratory confirmed: Serologically confirmed HIV status through point-of-care rapid test or laboratory confirmation. - Self-report:  Self-reported HIV status. | Methods employed to assess HIV prevalence.  One of:   - Laboratory confirmed: Presence of ART metabolites confirmed through laboratory testing. - VLS:  For studies reporting the proportion of the population that was virally suppressed, rather than on treatment, this proportion was divided by 0.9 to approximate ART coverage. - Self-report:  Self-reported ART status |
| Area | Surveillance area (e.g. city, district) | Surveillance area (e.g. city, district) | Surveillance area (e.g. city, district) |
| Age group | Study reported age group | Study reported age group | Study reported age group |
| Central estimate | Population size estimate count | HIV prevalence | ART coverage |
| Lower estimate | Lower bound: Population size estimate count | Lower bound: HIV prevalence | Lower bound: ART coverage |
| Upper estimate | Upper bound: Population size estimate | Upper bound: HIV prevalence | Upper bound: ART coverage |
| Denominator | N/A | Denominator | Tested HIV positive denominator |
| Source | Study reference | Study reference | Study reference |

FSW = Female sex workers; MSM = Men who have sex with men; PWID = People who inject drugs; TGW = Transgender women; TGM = Transgender men; NSUM = Network Scaleup Model; SS-PSE = Successive Sampling-Population Size Estimation; ART = Antiretroviral Therapy; VLS = Viral load suppression.

### Supplementary Table S3: National availability of population size, HIV prevalence and ART coverage data by key population.

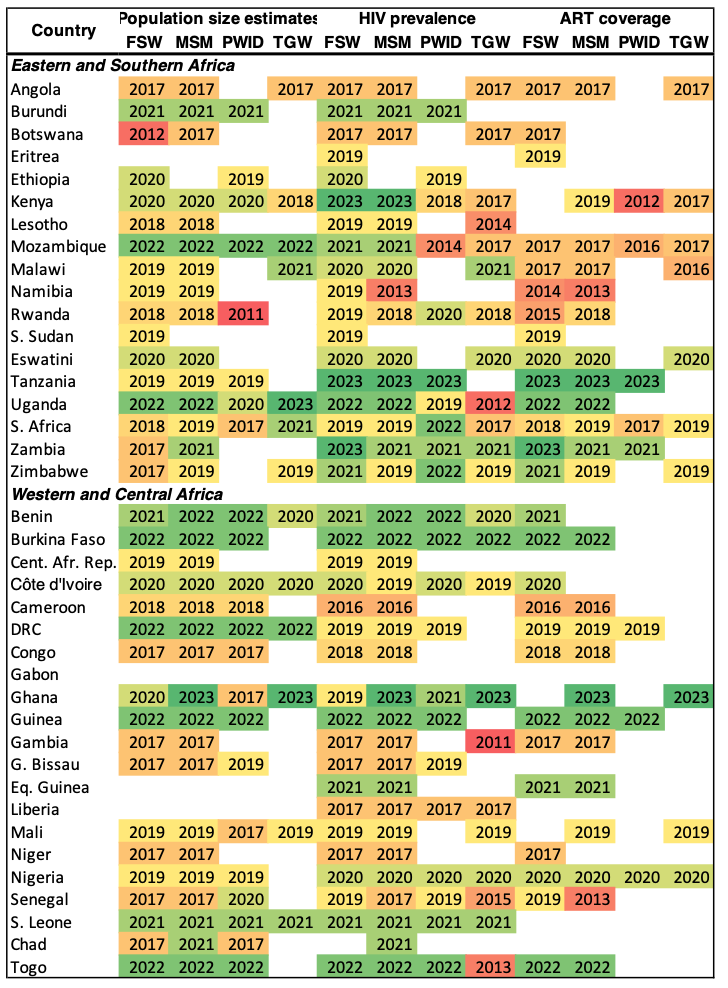

Cells are coloured by year with most recent in green, and oldest in red. FSW: female sex workers; MSM: men who have sex with men; PWID: people who inject drugs; TGW: transgender women

### Supplementary Table S4: Population size estimate method regression results

| Fixed effect category | **PSE method fixed effects (log odds ratios; 95% CI)** | | | |
| --- | --- | --- | --- | --- |
|  | **FSW** | **MSM** | **PWID** | **TGW** |
| Empirical | 0 (Reference category) | | | |
| PLACE/Mapping | **-0.55 (-0.92, -0.19)** | **-1.33 (-1.77, -0.90)** | **-1.16 (-1.68, -0.63)** | -1.32 (-2.66, 0.10) |

| **Fixed effect category** | **PSE method** | **PSE method random effects (log odds ratios; 95% CI)** | | | |
| --- | --- | --- | --- | --- | --- |
|  |  | **FSW** | **MSM** | **PWID** | **TGW** |
| Empirical | 2S-CRC | -0.04 (-0.29, 0.20) | -0.35 (-0.69, -0.07) | -0.03 (-0.41, 0.35) | -0.07 (-0.59, 0.35) |
| Empirical | SS-PSE | -0.01 (-0.31, 0.29) | -0.02 (-0.37, 0.28) | -0.10 (-0.57, 0.29) | -0.09 (-0.68, 0.36) |
| Empirical | MM - mixture | -0.01 (-0.39, 0.37) | 0.11 (-0.29, 0.56) | 0.11 (-0.32, 0.67) | *No observations* |
| Empirical | MM - empirical | -0.05 (-0.53, 0.35) | 0.12 (-0.35, 0.70) | -0.05 (-0.74, 0.51) | 0.17 (-0.35, 1.07) |
| Empirical | 3S-CRC | 0.09 (-0.19, 0.42) | 0 (-0.40, 0.38) | 0.07 (-0.33, 0.55) | -0.02 (-0.6, 0.49) |
| Empirical | NSUM | 0.02 (-0.42, 0.48) | 0.11 (-0.40, 0.76) | *No observations* | *No observations* |
| PLACE/Mapping | PLACE/Mapping |  |  |  |  |

Log odds ratios for population size estimates by methods. Two fixed effect categories were estimated: Empirical and PLACE/Mapping. Random effects were estimated for empirical methods (Two and three source CRC (2S- and 3S-CRC), network scale-up (NSUM), successive sampling population size estimation (SS-PSE), and average estimates from multiple empirical methods and estimates derived from a mixture of empirical and non-empirical methods (Multiple methods – mixed)). See Supplementary Figure S3 for a graphical representation of population size estimate method effects.

Each key population estimated in separate regression models, but results presented together for ease of comparability. PLACE: Priorities for Local AIDS Control Efforts; MM: Multiple methods; FSW: female sex workers; MSM: men who have sex with men; PWID: people who inject drugs; TGW: transgender women

### Supplementary Table S5: **HIV prevalence model regression results among female sex workers.**

| **Parameter** | **Log odds ratio (95%CI)** |
| --- | --- |
| Intercept | 1.36 (0.86, 1.88) |
| Logit total population prevalence | 0.71 (0.59, 0.84) |
| Region: ESA | 0 (Reference) |
| WCA | -2.79 (-3.62, -1.96) |
| Logit total population prevalence x region interaction | |
| Region: ESA | 0 (Reference) |
| WCA | -0.50 (-0.68, -0.34) |
| **Random effects** | **Precision (95% CI)** |
| National spatial effect | 10.56 (2.98, 41.91) |
| Provincial spatial effect | 4.75 (2.91, 7.79) |
| Study [subnational observation] | 7.13 (4.15, 12.15) |
| Study [national observation] | 10.99 (3.13, 43.4) |

ESA: Eastern and Southern Africa; WCA: Western and Central Africa

### Supplementary Table S6: **HIV prevalence model regression results among people who inject drugs.**

| **Parameter** | **Log odds ratio (95%CI)** |
| --- | --- |
| Intercept | -0.42 (-1.53, 0.64) |
| Logit total population prevalence | 0.29 (0.03, 0.56) |
| Region: ESA | 0 (Reference) |
| WCA | -2.1 (-4.08, -0.04) |
| Logit total population prevalence x region interaction | |
| Region: ESA | 0 (Reference) |
| WCA | -0.17 (-0.57, 0.24) |
| **Random effects** | **Precision (95% CI)** |
| National spatial effect | 6.19 (1.41, 35.69) |
| Provincial spatial effect | 4.79 (1.93, 11.91) |
| Study [subnational observation] | 2.42 (1.34, 4.38) |
| Study [national observation] | 6.69 (1.24, 39.61) |

ESA: Eastern and Southern Africa; WCA: Western and Central Africa

### Supplementary Table S7: **HIV prevalence model regression results among men who have sex with men and transgender women**

| **Parameter** | **Log odds ratio (95%CI)** |
| --- | --- |
| Intercept | -0.42 (-1.26, 0.4) |
| Logit total population prevalence | 0.55 (0.35, 0.76) |
| Region: ESA | 0 (Reference) |
| WCA | -0.15 (-1.49, 1.2) |
| Population: MSM | 0 (Reference) |
| TGW | 0.06 (-0.44, 0.57) |
| Logit total population prevalence x region interaction | |
| Region: ESA | 0 (Reference) |
| WCA | -0.25 (-0.53, 0.02) |
| Logit total population prevalence x KP interaction | |
| KP: MSM | 0 (Reference) |
| TGW | -0.03 (-0.25, 0.19) |
| **Random effect** | **Precision (95% CI)** |
| National spatial effect | 8.98 (2.11, 41.81) |
| Provincial spatial effect | 2.03 (1.31, 3.2) |
| Study [subnational observation] | 4.45 (2.7, 7.66) |
| Study [national observation] | 10.83 (2.54, 51.15) |

MSM-TGW were modelled together. MSM: men who have sex with men, TGW: transgender women, KP: key population; ESA: Eastern and Southern Africa; WCA: Western and Central Africa

| **Parameter** | | **Diagnostically-confirmed data only^a^**  **Log odds ratio (95%CI)** | **Diagnostically confirmed + self-report data**  **Log odds ratio (95%CI)** |
| --- | --- | --- | --- |
| Fixed effects | |  |  |
| Intercept | | -0.34 (-0.81, 0.09) | -0.35 (-0.82, 0.09) |
| Logit total population ART coverage | | 0.51 (0.23, 0.78) | 0.29 (0.05, 0.51) |
| Method: Diagnostically confirmed | |  | 0 (Reference category) |
| Self-report | |  | -0.31 (-0.74, 0.14) |
| Random effects | |  |  |
| Key population intercepts | | | |
|  | FSW | 0.03 (-0.4, 0.5) | 0.07 (-0.3, 0.48) |
|  | MSM | 0.25 (-0.14, 0.74) | 0 (-0.37, 0.39) |
|  | PWID | -0.15 (-0.71, 0.31) | -0.01 (-0.47, 0.46) |
|  | TGW | -0.12 (-0.65, 0.35) | -0.06 (-0.53, 0.35) |
| Key population random slopes | | | |
|  | FSW | 0.24 (-0.01, 0.57) | 0.11 (-0.08, 0.36) |
|  | MSM | -0.12 (-0.42, 0.15) | -0.06 (-0.29, 0.15) |
|  | PWID | 0.03 (-0.28, 0.38) | 0.01 (-0.25, 0.3) |
|  | TGW | -0.15 (-0.52, 0.13) | -0.06 (-0.34, 0.16) |
| **Parameter** |  | **Precision (95% CI)** |  |
| Key population intercepts | | 11.17 (2.25, 56.12) | 15.73 (2.95, 92.04) |
| Key population random slopes | | 29.55 (7.03, 134.35) | 57.19 (11.79, 321.17) |
| National spatial model | | 52.72 (7.87, 368.56) | 19.6 (3.22, 141.15) |
| Provincial spatial model | | 39.74 (7.72, 278.85) | 53.44 (10.4, 361.33) |
| Study [subnational observation] | | 6.36 (2.28, 20.35) | 1.42 (0.81, 2.61) |
| Study [national observation] | | 8.11 (1.53, 50.01) | 4.09 (1, 15.4) |

### Supplementary Table S8: ART coverage model regression results.

^a^ Diagnostically-confirmed data includes viral load suppression data, converted to an estimate of ART coverage by assuming 90% of those on ART are virally suppressed, and laboratory-confirmed ART status data from antiretroviral drug metabolite testing.

FSW: female sex workers, MSM: men who have sex with men, PWID: people who inject drugs; TGW: transgender women

### Supplementary Table S9: Country-specific KPSE proportions (%) for female sex workers

|  |  | **Total population urban proportion (%)^1^** | **Urban KPSE**  **(%; 95% CI)** | **Total KPSE**  **(%; 95% CI)** |
| --- | --- | --- | --- | --- |
| ESA | Angola | 67 | 1.95 (1.04, 3.71) | 1.76 (0.91, 3.31) |
|  | Burundi | 14 | 1.82 (0.90, 3.53) | 1.20 (0.52, 2.54) |
|  | Botswana | 71 | 2.14 (1.13, 5.01) | 1.91 (1.02, 4.53) |
|  | Eritrea | 41 | 1.69 (0.30, 9.22) | 1.29 (0.24, 7.05) |
|  | Ethiopia | 22 | 1.70 (0.88, 3.27) | 1.18 (0.55, 2.42) |
|  | Kenya | 28 | 2.24 (1.33, 3.89) | 1.62 (0.86, 2.94) |
|  | Lesotho | 29 | 1.51 (0.59, 3.88) | 1.09 (0.40, 2.89) |
|  | Mozambique | 37 | 1.60 (0.97, 2.50) | 1.21 (0.68, 2.12) |
|  | Malawi | 17 | 1.98 (1.18, 3.40) | 1.34 (0.66, 2.57) |
|  | Namibia | 52 | 2.16 (1.17, 3.96) | 1.77 (0.95, 3.35) |
|  | Rwanda | 17 | 1.60 (0.70, 3.43) | 1.08 (0.42, 2.53) |
|  | S. Sudan | 20 | 2.51 (1.33, 5.09) | 1.77 (0.86, 3.66) |
|  | Eswatini | 24 | 2.03 (0.84, 4.79) | 1.41 (0.57, 3.52) |
|  | Tanzania | 35 | 1.52 (0.92, 2.47) | 1.13 (0.66, 1.93) |
|  | Uganda | 25 | 1.70 (1.03, 2.91) | 1.21 (0.66, 2.20) |
|  | S. Africa | 67 | 1.38 (0.69, 2.41) | 1.20 (0.60, 2.15) |
|  | Zambia | 45 | 1.78 (1.10, 2.86) | 1.42 (0.80, 2.32) |
|  | Zimbabwe | 32 | 1.61 (0.85, 3.10) | 1.18 (0.58, 2.46) |
|  | ***Pooled region*** |  | **1.76 (1.31, 2.38)** | **1.33 (0.89, 1.99)** |
| WCA | Benin | 48 | 1.40 (0.75, 2.84) | 1.13 (0.60, 2.34) |
|  | Burkina Faso | 31 | 0.94 (0.51, 1.64) | 0.69 (0.37, 1.28) |
|  | Cent. Afr. Rep. | 42 | 2.65 (1.42, 5.82) | 2.02 (1.09, 4.54) |
|  | Côte d'Ivoire | 52 | 1.12 (0.61, 2.31) | 0.92 (0.48, 1.92) |
|  | Cameroon | 58 | 2.10 (1.20, 3.76) | 1.77 (1.01, 3.28) |
|  | Dem. Rep. Congo | 46 | 1.81 (1.15, 2.81) | 1.41 (0.86, 2.35) |
|  | Congo | 68 | 1.78 (0.90, 3.40) | 1.56 (0.79, 3.04) |
|  | Gabon | 90 | 1.98 (0.64, 5.48) | 1.92 (0.62, 5.38) |
|  | Ghana | 57 | 0.45 (0.22, 0.92) | 0.38 (0.18, 0.78) |
|  | Guinea | 37 | 1.39 (0.78, 2.56) | 1.07 (0.56, 1.99) |
|  | Gambia | 63 | 1.10 (0.32, 4.44) | 0.95 (0.26, 3.82) |
|  | G. Bissau | 44 | 1.29 (0.47, 3.49) | 1.01 (0.37, 2.84) |
|  | Eq. Guinea | 73 | 1.96 (0.57, 7.87) | 1.77 (0.49, 7.13) |
|  | Liberia | 52 | 1.30 (0.50, 4.12) | 1.07 (0.39, 3.42) |
|  | Mali | 44 | 1.04 (0.58, 1.85) | 0.81 (0.44, 1.55) |
|  | Niger | 17 | 1.57 (0.81, 3.37) | 1.05 (0.46, 2.42) |
|  | Nigeria | 52 | 2.02 (1.18, 3.60) | 1.66 (0.92, 2.85) |
|  | Senegal | 48 | 0.99 (0.42, 1.99) | 0.80 (0.33, 1.65) |
|  | S. Leone | 43 | 1.40 (0.58, 3.38) | 1.10 (0.44, 2.86) |
|  | Chad | 24 | 3.11 (1.40, 7.98) | 2.20 (0.91, 5.89) |
|  | Togo | 43 | 1.41 (0.73, 2.69) | 1.12 (0.57, 2.28) |
|  | ***Pooled region*** |  | **1.73 (1.22, 2.55)** | **1.39 (0.92, 2.05)** |
| **SSA** | ***Pooled region*** |  | **1.76 (1.33, 2.30)** | **1.38 (0.96, 1.93)** |

1: Extracted from UN Population Division World Urbanisation Prospects. ESA: Eastern and Southern Africa; WCA: Western and Central Africa; SSA: sub-Saharan Africa; KPSE: Key population size estimate

### Supplementary Table S10: Country-specific KPSE proportions (%) for men who have sex with men

|  |  | **Total population urban proportion (%)^1^** | **Urban KPSE**  **(%; 95% CI)** | **Total KPSE**  **(%; 95% CI)** |
| --- | --- | --- | --- | --- |
| ESA | Angola | 67 | 0.97 (0.58, 1.73) | 0.89 (0.51, 1.54) |
|  | Burundi | 14 | 0.95 (0.55, 1.71) | 0.64 (0.32, 1.23) |
|  | Botswana | 71 | 0.94 (0.57, 1.56) | 0.84 (0.50, 1.39) |
|  | Eritrea | 41 | 0.85 (0.30, 2.48) | 0.66 (0.22, 1.89) |
|  | Ethiopia | 22 | 0.84 (0.39, 1.70) | 0.59 (0.26, 1.22) |
|  | Kenya | 28 | 0.80 (0.47, 1.29) | 0.59 (0.30, 1.02) |
|  | Lesotho | 29 | 0.95 (0.45, 2.14) | 0.70 (0.31, 1.61) |
|  | Mozambique | 37 | 0.98 (0.65, 1.57) | 0.75 (0.44, 1.24) |
|  | Malawi | 17 | 0.99 (0.60, 1.73) | 0.68 (0.34, 1.25) |
|  | Namibia | 52 | 1.02 (0.63, 1.84) | 0.86 (0.51, 1.56) |
|  | Rwanda | 17 | 0.93 (0.54, 1.73) | 0.64 (0.33, 1.24) |
|  | S. Sudan | 20 | 0.87 (0.50, 1.49) | 0.61 (0.33, 1.19) |
|  | Eswatini | 24 | 0.98 (0.53, 1.94) | 0.69 (0.34, 1.40) |
|  | Tanzania | 35 | 0.94 (0.60, 1.56) | 0.72 (0.41, 1.17) |
|  | Uganda | 25 | 0.92 (0.59, 1.49) | 0.66 (0.36, 1.14) |
|  | S. Africa | 67 | 0.88 (0.54, 1.38) | 0.79 (0.48, 1.24) |
|  | Zambia | 45 | 0.96 (0.62, 1.52) | 0.77 (0.46, 1.24) |
|  | Zimbabwe | 32 | 0.91 (0.54, 1.54) | 0.69 (0.38, 1.23) |
|  | ***Pooled region*** |  | **0.92 (0.65, 1.36)** | **0.70 (0.43, 1.08)** |
| WCA | Benin | 48 | 0.78 (0.46, 1.36) | 0.63 (0.36, 1.13) |
|  | Burkina Faso | 31 | 0.70 (0.41, 1.11) | 0.54 (0.29, 0.88) |
|  | Cent. Afr. Rep. | 42 | 0.94 (0.57, 1.66) | 0.74 (0.41, 1.28) |
|  | Côte d'Ivoire | 52 | 0.74 (0.46, 1.17) | 0.60 (0.37, 0.97) |
|  | Cameroon | 58 | 0.98 (0.58, 1.75) | 0.84 (0.49, 1.51) |
|  | Dem. Rep. Congo | 46 | 0.93 (0.59, 1.47) | 0.74 (0.45, 1.18) |
|  | Congo | 68 | 0.95 (0.54, 1.74) | 0.83 (0.47, 1.52) |
|  | Gabon | 90 | 0.98 (0.49, 2.26) | 0.94 (0.47, 2.15) |
|  | Ghana | 57 | 0.78 (0.47, 1.30) | 0.67 (0.39, 1.10) |
|  | Guinea | 37 | 0.64 (0.36, 1.08) | 0.49 (0.26, 0.87) |
|  | Gambia | 63 | 0.69 (0.25, 1.80) | 0.61 (0.23, 1.59) |
|  | G. Bissau | 44 | 0.66 (0.31, 1.27) | 0.54 (0.23, 1.06) |
|  | Eq. Guinea | 73 | 0.96 (0.41, 2.32) | 0.87 (0.37, 2.12) |
|  | Liberia | 52 | 0.71 (0.39, 1.37) | 0.58 (0.31, 1.12) |
|  | Mali | 44 | 0.75 (0.46, 1.22) | 0.59 (0.34, 1.01) |
|  | Niger | 17 | 0.83 (0.50, 1.39) | 0.56 (0.28, 1.06) |
|  | Nigeria | 52 | 0.89 (0.56, 1.51) | 0.73 (0.44, 1.25) |
|  | Senegal | 48 | 0.66 (0.33, 1.20) | 0.54 (0.26, 0.99) |
|  | S. Leone | 43 | 0.79 (0.38, 1.58) | 0.62 (0.29, 1.27) |
|  | Chad | 24 | 1.00 (0.61, 1.81) | 0.72 (0.39, 1.35) |
|  | Togo | 43 | 0.82 (0.48, 1.41) | 0.65 (0.38, 1.11) |
|  | ***Pooled region*** |  | **0.87 (0.62, 1.29)** | **0.70 (0.47, 1.06)** |
| **SSA** | ***Pooled region*** |  | **0.90 (0.66, 1.24)** | **0.71 (0.48, 1.03)** |

1: Extracted from UN Population Division World Urbanisation Prospects. ESA: Eastern and Southern Africa; WCA: Western and Central Africa; SSA: sub-Saharan Africa; KPSE: Key population size estimate

### Supplementary Table S11: Country-specific KPSE proportions (%) for people who inject drugs

|  |  | **Total population urban proportion (%)^1^** | **Urban KPSE**  **(%; 95% CI)** | **Total KPSE**  **(%; 95% CI)** |
| --- | --- | --- | --- | --- |
| ESA | Angola | 67 | 0.32 (0.16, 0.59) | 0.29 (0.14, 0.55) |
|  | Burundi | 14 | 0.35 (0.19, 0.71) | 0.24 (0.11, 0.50) |
|  | Botswana | 71 | 0.32 (0.14, 0.64) | 0.28 (0.13, 0.58) |
|  | Eritrea | 41 | 0.35 (0.10, 1.29) | 0.27 (0.07, 1.00) |
|  | Ethiopia | 22 | 0.34 (0.17, 0.80) | 0.24 (0.11, 0.56) |
|  | Kenya | 28 | 0.35 (0.21, 0.62) | 0.26 (0.14, 0.46) |
|  | Lesotho | 29 | 0.30 (0.09, 0.84) | 0.22 (0.07, 0.59) |
|  | Mozambique | 37 | 0.32 (0.17, 0.55) | 0.24 (0.12, 0.44) |
|  | Malawi | 17 | 0.33 (0.16, 0.66) | 0.22 (0.10, 0.49) |
|  | Namibia | 52 | 0.31 (0.15, 0.62) | 0.26 (0.12, 0.54) |
|  | Rwanda | 17 | 0.34 (0.18, 0.66) | 0.23 (0.11, 0.49) |
|  | S. Sudan | 20 | 0.35 (0.19, 0.65) | 0.24 (0.12, 0.48) |
|  | Eswatini | 24 | 0.31 (0.13, 0.66) | 0.22 (0.09, 0.48) |
|  | Tanzania | 35 | 0.34 (0.21, 0.57) | 0.25 (0.14, 0.45) |
|  | Uganda | 25 | 0.34 (0.20, 0.60) | 0.24 (0.13, 0.45) |
|  | S. Africa | 67 | 0.30 (0.14, 0.56) | 0.27 (0.12, 0.49) |
|  | Zambia | 45 | 0.32 (0.17, 0.57) | 0.26 (0.13, 0.46) |
|  | Zimbabwe | 32 | 0.31 (0.14, 0.67) | 0.24 (0.10, 0.50) |
|  | ***Pooled region*** |  | **0.34 (0.21, 0.53)** | **0.26 (0.15, 0.44)** |
| WCA | Benin | 48 | 0.36 (0.21, 0.64) | 0.29 (0.16, 0.55) |
|  | Burkina Faso | 31 | 0.36 (0.21, 0.60) | 0.27 (0.15, 0.48) |
|  | Cent. Afr. Rep. | 42 | 0.35 (0.19, 0.63) | 0.27 (0.14, 0.51) |
|  | Côte d'Ivoire | 52 | 0.37 (0.19, 0.70) | 0.30 (0.15, 0.59) |
|  | Cameroon | 58 | 0.34 (0.19, 0.63) | 0.28 (0.15, 0.53) |
|  | Dem. Rep. Congo | 46 | 0.33 (0.20, 0.51) | 0.26 (0.15, 0.42) |
|  | Congo | 68 | 0.32 (0.16, 0.60) | 0.28 (0.14, 0.53) |
|  | Gabon | 90 | 0.34 (0.14, 0.72) | 0.33 (0.13, 0.69) |
|  | Ghana | 57 | 0.37 (0.18, 0.71) | 0.31 (0.14, 0.62) |
|  | Guinea | 37 | 0.37 (0.18, 0.67) | 0.28 (0.13, 0.53) |
|  | Gambia | 63 | 0.37 (0.11, 1.09) | 0.32 (0.10, 0.97) |
|  | G. Bissau | 44 | 0.39 (0.17, 0.93) | 0.31 (0.14, 0.77) |
|  | Eq. Guinea | 73 | 0.34 (0.13, 0.85) | 0.31 (0.12, 0.78) |
|  | Liberia | 52 | 0.40 (0.18, 0.84) | 0.33 (0.14, 0.70) |
|  | Mali | 44 | 0.39 (0.22, 0.71) | 0.31 (0.17, 0.58) |
|  | Niger | 17 | 0.39 (0.22, 0.70) | 0.26 (0.13, 0.53) |
|  | Nigeria | 52 | 0.42 (0.24, 0.79) | 0.35 (0.20, 0.66) |
|  | Senegal | 48 | 0.36 (0.16, 0.69) | 0.29 (0.13, 0.60) |
|  | S. Leone | 43 | 0.48 (0.24, 1.02) | 0.38 (0.18, 0.85) |
|  | Chad | 24 | 0.40 (0.22, 0.86) | 0.29 (0.15, 0.64) |
|  | Togo | 43 | 0.40 (0.23, 0.87) | 0.32 (0.17, 0.71) |
|  | ***Pooled region*** |  | **0.39 (0.26, 0.61)** | **0.32 (0.20, 0.51)** |
| **SSA** | ***Pooled region*** |  | **0.37 (0.25, 0.55)** | **0.29 (0.18, 0.46)** |

1: Extracted from UN Population Division World Urbanisation Prospects. ESA: Eastern and Southern Africa; WCA: Western and Central Africa; SSA: sub-Saharan Africa; KPSE: Key population size estimate

### Supplementary Table S12: Country-specific KPSE proportions (%) for transgender women

|  |  | **Total population urban proportion (%)^1^** | **Urban KPSE**  **(%; 95% CI)** | **Total KPSE**  **(%; 95% CI)** |
| --- | --- | --- | --- | --- |
| ESA | Angola | 67 | 0.11 (0.04, 0.30) | 0.10 (0.04, 0.27) |
|  | Burundi | 14 | 0.12 (0.04, 0.40) | 0.08 (0.02, 0.28) |
|  | Botswana | 71 | 0.12 (0.05, 0.43) | 0.11 (0.04, 0.38) |
|  | Eritrea | 41 | 0.10 (0.02, 0.62) | 0.08 (0.01, 0.47) |
|  | Ethiopia | 22 | 0.11 (0.03, 0.38) | 0.07 (0.02, 0.27) |
|  | Kenya | 28 | 0.10 (0.04, 0.30) | 0.07 (0.03, 0.23) |
|  | Lesotho | 29 | 0.13 (0.03, 0.76) | 0.09 (0.02, 0.58) |
|  | Mozambique | 37 | 0.12 (0.05, 0.30) | 0.09 (0.03, 0.22) |
|  | Malawi | 17 | 0.11 (0.05, 0.35) | 0.08 (0.03, 0.23) |
|  | Namibia | 52 | 0.12 (0.05, 0.42) | 0.10 (0.04, 0.36) |
|  | Rwanda | 17 | 0.11 (0.04, 0.36) | 0.08 (0.03, 0.25) |
|  | S. Sudan | 20 | 0.11 (0.04, 0.34) | 0.08 (0.03, 0.26) |
|  | Eswatini | 24 | 0.12 (0.04, 0.51) | 0.09 (0.03, 0.38) |
|  | Tanzania | 35 | 0.11 (0.05, 0.31) | 0.08 (0.03, 0.24) |
|  | Uganda | 25 | 0.11 (0.04, 0.32) | 0.08 (0.03, 0.23) |
|  | S. Africa | 67 | 0.13 (0.05, 0.43) | 0.12 (0.04, 0.38) |
|  | Zambia | 45 | 0.12 (0.05, 0.31) | 0.09 (0.04, 0.26) |
|  | Zimbabwe | 32 | 0.12 (0.05, 0.30) | 0.09 (0.03, 0.23) |
|  | ***Pooled region*** |  | **0.12 (0.05, 0.31)** | **0.09 (0.04, 0.23)** |
| WCA | Benin | 48 | 0.09 (0.03, 0.21) | 0.07 (0.03, 0.17) |
|  | Burkina Faso | 31 | 0.07 (0.02, 0.17) | 0.05 (0.02, 0.13) |
|  | Cent. Afr. Rep. | 42 | 0.10 (0.04, 0.33) | 0.08 (0.03, 0.27) |
|  | Côte d'Ivoire | 52 | 0.07 (0.02, 0.17) | 0.06 (0.02, 0.14) |
|  | Cameroon | 58 | 0.10 (0.03, 0.32) | 0.09 (0.03, 0.27) |
|  | Dem. Rep. Congo | 46 | 0.12 (0.05, 0.32) | 0.10 (0.04, 0.25) |
|  | Congo | 68 | 0.11 (0.04, 0.35) | 0.10 (0.03, 0.31) |
|  | Gabon | 90 | 0.10 (0.03, 0.40) | 0.10 (0.03, 0.38) |
|  | Ghana | 57 | 0.07 (0.02, 0.17) | 0.06 (0.02, 0.15) |
|  | Guinea | 37 | 0.07 (0.02, 0.18) | 0.06 (0.02, 0.14) |
|  | Gambia | 63 | 0.07 (0.01, 0.36) | 0.06 (0.01, 0.31) |
|  | G. Bissau | 44 | 0.07 (0.02, 0.25) | 0.06 (0.01, 0.21) |
|  | Eq. Guinea | 73 | 0.10 (0.02, 0.50) | 0.09 (0.02, 0.44) |
|  | Liberia | 52 | 0.08 (0.02, 0.23) | 0.06 (0.02, 0.18) |
|  | Mali | 44 | 0.06 (0.02, 0.15) | 0.05 (0.02, 0.13) |
|  | Niger | 17 | 0.08 (0.03, 0.21) | 0.05 (0.02, 0.15) |
|  | Nigeria | 52 | 0.09 (0.03, 0.27) | 0.07 (0.02, 0.22) |
|  | Senegal | 48 | 0.07 (0.02, 0.21) | 0.06 (0.02, 0.17) |
|  | S. Leone | 43 | 0.09 (0.03, 0.29) | 0.07 (0.02, 0.24) |
|  | Chad | 24 | 0.09 (0.03, 0.29) | 0.06 (0.02, 0.21) |
|  | Togo | 43 | 0.08 (0.02, 0.22) | 0.06 (0.02, 0.17) |
|  | ***Pooled region*** |  | **0.09 (0.04, 0.21)** | **0.08 (0.03, 0.17)** |
| **SSA** | ***Pooled region*** |  | **0.11 (0.05, 0.23)** | **0.08 (0.04, 0.19)** |

1: Extracted from UN Population Division World Urbanisation Prospects. ESA: Eastern and Southern Africa; WCA: Western and Central Africa; SSA: sub-Saharan Africa; KPSE: Key population size estimate

### Supplementary Table S13: Country-specific KP HIV prevalence estimates

|  |  | **HIV Prevalence (%) (95% CI)** | | | |
| --- | --- | --- | --- | --- | --- |
| **Region** | **Country** | **FSW** | **MSM** | **PWID** | **TGW** |
| ESA | Angola | 13 (7, 21) | 5 (2, 10) | 14 (6, 36) | 8 (4, 18) |
|  | Burundi | 13 (9, 20) | 4 (2, 8) | 9 (4, 19) | 6 (3, 15) |
|  | Botswana | 50 (40, 61) | 17 (10, 26) | 26 (11, 46) | 25 (15, 38) |
|  | Eritrea | 13 (6, 27) | 4 (1, 25) | 12 (2, 45) | 7 (1, 41) |
|  | Ethiopia | 14 (10, 21) | 5 (2, 16) | 12 (5, 28) | 9 (2, 28) |
|  | Kenya | 26 (17, 35) | 12 (8, 19) | 22 (14, 33) | 20 (12, 30) |
|  | Lesotho | 57 (45, 69) | 29 (20, 40) | 27 (7, 61) | 39 (27, 54) |
|  | Mozambique | 35 (27, 44) | 14 (9, 20) | 26 (15, 41) | 21 (14, 30) |
|  | Malawi | 43 (30, 55) | 13 (9, 18) | 21 (9, 40) | 20 (14, 26) |
|  | Namibia | 36 (26, 46) | 14 (8, 25) | 22 (9, 42) | 21 (12, 35) |
|  | Rwanda | 28 (21, 36) | 6 (4, 11) | 14 (7, 26) | 10 (5, 19) |
|  | S. Sudan | 22 (14, 31) | 7 (3, 16) | 18 (9, 35) | 12 (5, 25) |
|  | Eswatini | 62 (47, 74) | 24 (15, 37) | 32 (13, 60) | 37 (23, 53) |
|  | Tanzania | 27 (21, 35) | 12 (8, 17) | 21 (15, 28) | 19 (12, 27) |
|  | Uganda | 33 (26, 42) | 10 (6, 19) | 26 (14, 44) | 16 (8, 27) |
|  | S. Africa | 59 (53, 65) | 33 (27, 39) | 27 (17, 43) | 45 (36, 55) |
|  | Zambia | 45 (38, 53) | 15 (9, 22) | 23 (12, 35) | 21 (13, 31) |
|  | Zimbabwe | 48 (39, 57) | 16 (9, 25) | 23 (11, 41) | 22 (14, 34) |
|  | ***Pooled region*** | **30 (25, 34)** | **13 (10, 17)** | **20 (15, 27)** | **20 (15, 29)** |
| WCA | Benin | 12 (9, 16) | 10 (6, 14) | 3 (2, 6) | 14 (9, 21) |
|  | Burkina Faso | 9 (7, 13) | 7 (5, 11) | 3 (1, 6) | 10 (7, 16) |
|  | Cent. Afr. Rep. | 13 (9, 20) | 16 (9, 25) | 5 (2, 14) | 20 (12, 32) |
|  | Côte d'Ivoire | 7 (5, 10) | 10 (7, 15) | 4 (2, 7) | 14 (9, 20) |
|  | Cameroon | 17 (11, 26) | 18 (12, 28) | 5 (2, 13) | 23 (15, 35) |
|  | Dem. Rep. Congo | 8 (6, 13) | 11 (6, 20) | 4 (2, 11) | 15 (8, 26) |
|  | Congo | 11 (7, 18) | 22 (13, 33) | 5 (2, 15) | 28 (17, 43) |
|  | Gabon | 12 (6, 24) | 16 (6, 34) | 5 (1, 16) | 23 (10, 46) |
|  | Ghana | 8 (5, 12) | 26 (19, 36) | 4 (2, 7) | 33 (25, 45) |
|  | Guinea | 9 (6, 14) | 10 (6, 16) | 5 (2, 10) | 13 (8, 21) |
|  | Gambia | 12 (6, 21) | 17 (10, 28) | 5 (1, 23) | 22 (12, 34) |
|  | G. Bissau | 15 (8, 25) | 17 (7, 34) | 6 (2, 14) | 21 (9, 40) |
|  | Eq. Guinea | 15 (6, 33) | 20 (6, 48) | 5 (1, 22) | 24 (8, 54) |
|  | Liberia | 10 (6, 16) | 15 (9, 24) | 5 (3, 10) | 19 (11, 31) |
|  | Mali | 10 (7, 16) | 11 (8, 16) | 4 (2, 10) | 15 (10, 21) |
|  | Niger | 10 (7, 15) | 6 (4, 11) | 3 (1, 7) | 8 (4, 15) |
|  | Nigeria | 14 (10, 18) | 14 (10, 19) | 4 (2, 7) | 18 (12, 26) |
|  | Senegal | 8 (5, 13) | 23 (15, 31) | 5 (2, 11) | 28 (19, 40) |
|  | S. Leone | 10 (5, 16) | 6 (3, 13) | 6 (3, 13) | 10 (5, 20) |
|  | Chad | 14 (8, 24) | 8 (4, 16) | 4 (2, 11) | 11 (6, 20) |
|  | Togo | 13 (9, 17) | 18 (13, 26) | 5 (3, 9) | 24 (16, 33) |
|  | ***Pooled region*** | **12 (10, 15)** | **14 (11, 17)** | **4 (3, 6)** | **18 (14, 23)** |
| **SSA** | ***Pooled region*** | **21 (18, 24)** | **14 (12, 16)** | **11 (8, 15)** | **19 (15, 25)** |

FSW: female sex workers; MSM: men who have sex with men; PWID: people who inject drugs; TGW: transgender women; ESA: Eastern and Southern Africa; WCA: Western and Central Africa; SSA: sub-Saharan Africa

### Supplementary Table S14: Country-specific KP ART coverage estimates

| **Region** | **Country** | **ART Coverage (%) (95% CI)** | | | |
| --- | --- | --- | --- | --- | --- |
|  |  | **FSW** | **MSM** | **PWID** | **TGW** |
| **ESA** | Angola | 47 (28, 68) | 50 (30, 71) | 36 (16, 61) | 48 (27, 70) |
|  | Burundi | 86 (70, 95) | 68 (44, 86) | 63 (36, 84) | 62 (35, 85) |
|  | Botswana | 100 (100, 100) | 67 (46, 83) | 64 (38, 84) | 93 (25, 100) |
|  | Eritrea | 53 (13, 87) | 49 (12, 85) | 40 (8, 81) | 41 (9, 81) |
|  | Ethiopia | 75 (53, 89) | 59 (31, 82) | 52 (23, 74) | 56 (26, 81) |
|  | Kenya | 81 (70, 88) | 61 (47, 73) | 55 (37, 72) | 53 (32, 72) |
|  | Lesotho | 74 (39, 92) | 53 (21, 81) | 46 (16, 76) | 47 (17, 80) |
|  | Mozambique | 70 (56, 81) | 57 (42, 71) | 51 (33, 69) | 49 (33, 65) |
|  | Malawi | 87 (70, 94) | 61 (40, 77) | 59 (33, 81) | 54 (29, 77) |
|  | Namibia | 90 (78, 95) | 66 (45, 83) | 62 (38, 83) | 63 (31, 85) |
|  | Rwanda | 88 (78, 95) | 74 (56, 87) | 71 (47, 87) | 65 (43, 85) |
|  | S. Sudan | 23 (11, 41) | 39 (21, 61) | 26 (11, 49) | 32 (14, 57) |
|  | Eswatini | 90 (73, 97) | 64 (37, 85) | 61 (30, 85) | 59 (24, 88) |
|  | Tanzania | 94 (88, 97) | 70 (54, 84) | 69 (46, 86) | 68 (37, 89) |
|  | Uganda | 80 (71, 87) | 64 (50, 75) | 57 (40, 74) | 57 (40, 74) |
|  | S. Africa | 61 (52, 71) | 48 (39, 58) | 39 (27, 51) | 44 (32, 56) |
|  | Zambia | 85 (77, 90) | 71 (57, 81) | 68 (50, 82) | 61 (40, 77) |
|  | Zimbabwe | 91 (85, 95) | 78 (64, 87) | 77 (58, 89) | 75 (36, 87) |
|  | ***Pooled region*** | 75 (68, 81) | 58 (49, 66) | 53 (40, 65) | 52 (37, 67) |
| **WCA** | Benin | 75 (51, 89) | 53 (28, 75) | 47 (21, 72) | 48 (21, 74) |
|  | Burkina Faso | 71 (50, 84) | 48 (29, 65) | 45 (24, 68) | 41 (20, 64) |
|  | Cent. Afr. Rep. | 44 (24, 65) | 48 (25, 71) | 36 (16, 60) | 42 (22, 66) |
|  | Côte d'Ivoire | 65 (48, 80) | 48 (30, 66) | 37 (21, 58) | 45 (27, 67) |
|  | Cameroon | 88 (72, 95) | 66 (41, 86) | 62 (35, 84) | 60 (31, 86) |
|  | Dem. Rep. Congo | 80 (71, 87) | 69 (56, 81) | 65 (46, 80) | 60 (42, 76) |
|  | Congo | 19 (9, 36) | 40 (22, 63) | 24 (9, 53) | 36 (14, 65) |
|  | Gabon | 47 (21, 72) | 58 (28, 81) | 49 (21, 76) | 45 (18, 73) |
|  | Ghana | 50 (33, 65) | 54 (39, 67) | 44 (27, 63) | 43 (27, 60) |
|  | Guinea | 52 (35, 68) | 39 (23, 54) | 28 (14, 48) | 38 (22, 57) |
|  | Gambia | 32 (10, 64) | 32 (11, 63) | 19 (4, 53) | 35 (11, 68) |
|  | G. Bissau | 58 (28, 84) | 42 (15, 74) | 32 (11, 65) | 43 (17, 76) |
|  | Eq. Guinea | 48 (23, 75) | 40 (13, 76) | 25 (7, 68) | 46 (17, 78) |
|  | Liberia | 63 (35, 84) | 47 (23, 74) | 38 (15, 67) | 43 (20, 72) |
|  | Mali | 38 (24, 54) | 42 (28, 58) | 31 (17, 49) | 37 (23, 54) |
|  | Niger | 78 (58, 90) | 47 (26, 68) | 37 (17, 61) | 51 (24, 77) |
|  | Nigeria | 69 (51, 82) | 59 (33, 76) | 51 (28, 70) | 50 (25, 72) |
|  | Senegal | 65 (36, 86) | 49 (21, 78) | 41 (16, 72) | 46 (19, 77) |
|  | S. Leone | 69 (37, 90) | 55 (26, 82) | 47 (20, 75) | 45 (18, 78) |
|  | Chad | 72 (49, 86) | 55 (28, 75) | 47 (21, 70) | 51 (25, 74) |
|  | Togo | 78 (54, 91) | 46 (22, 71) | 36 (15, 63) | 50 (21, 78) |
|  | ***Pooled region*** | 70 (57, 79) | 57 (42, 68) | 49 (33, 62) | 50 (34, 66) |
| **SSA** | ***Regional Average*** | 73 (67, 79) | 57 (47, 66) | 52 (39, 63) | 52 (36, 64) |

FSW: female sex workers; MSM: men who have sex with men; PWID: people who inject drugs; TGW: transgender women; ESA: Eastern and Southern Africa; WCA: Western and Central Africa; SSA: sub-Saharan Africa

### Supplementary Table S15: Country-specific estimates of the proportion of all 15-49 PLHIV among key populations

| **Region** | **Country** | **15-49 PLHIV proportion (%) (95% CI)** | | | |
| --- | --- | --- | --- | --- | --- |
|  |  | **FSW** | **MSM** | **PWID** | **TGW** |
| **ESA** | Angola | 8.3 (3.2, 18.6) | 1.5 (0.6, 3.6) | 1.4 (0.4, 4.2) | 0.3 (0.1, 1.1) |
|  | Burundi | 2.9 (1.5, 7.3) | 0.4 (0.2, 0.9) | 0.2 (0.1, 0.5) | 0.1 (0.0, 0.3) |
|  | Botswana | 8.7 (3.7, 19.7) | 1.4 (0.6, 3.4) | 1.2 (0.4, 3.4) | 0.3 (0.1, 1.3) |
|  | Eritrea | 19.6 (2.8, 100.0) | 3.1 (0.3, 25.5) | 3.4 (0.4, 24.4) | 0.6 (0.0, 7.9) |
|  | Ethiopia | 1.6 (0.6, 4.3) | 0.3 (0.1, 0.8) | 0.1 (0.0, 0.4) | 0.1 (0.0, 0.3) |
|  | Kenya | 12.7 (5.5, 30.1) | 2.4 (0.6, 9.0) | 2.2 (0.7, 7.4) | 0.5 (0.1, 3.1) |
|  | Lesotho | 5.9 (2.9, 12.1) | 1.0 (0.5, 2.0) | 0.8 (0.4, 1.7) | 0.2 (0.1, 0.7) |
|  | Mozambique | 1.9 (0.7, 5.3) | 0.6 (0.2, 1.6) | 0.2 (0.0, 0.6) | 0.1 (0.0, 0.7) |
|  | Malawi | 4.0 (1.9, 8.0) | 0.6 (0.3, 1.2) | 0.3 (0.1, 0.8) | 0.1 (0.0, 0.4) |
|  | Namibia | 1.8 (1.0, 3.3) | 0.4 (0.2, 0.8) | 0.3 (0.1, 0.6) | 0.1 (0.0, 0.2) |
|  | Rwanda | 2.9 (1.4, 5.7) | 0.5 (0.3, 1.2) | 0.3 (0.1, 0.7) | 0.1 (0.0, 0.4) |
|  | S. Sudan | 7.0 (2.6, 16.8) | 0.9 (0.4, 2.2) | 0.7 (0.3, 2.0) | 0.2 (0.1, 0.6) |
|  | Eswatini | 1.9 (0.9, 3.6) | 0.7 (0.4, 1.2) | 0.2 (0.1, 0.4) | 0.1 (0.1, 0.5) |
|  | Tanzania | 6.9 (2.9, 16.1) | 0.8 (0.3, 2.3) | 0.8 (0.3, 2.1) | 0.2 (0.0, 0.6) |
|  | Uganda | 4.0 (2.1, 7.1) | 1.0 (0.5, 1.9) | 0.7 (0.3, 1.3) | 0.2 (0.1, 0.7) |
|  | S. Africa | 4.4 (2.2, 8.6) | 0.7 (0.3, 1.6) | 0.7 (0.3, 1.5) | 0.1 (0.0, 0.5) |
|  | Zambia | 3.0 (1.6, 5.3) | 0.5 (0.3, 1.0) | 0.3 (0.1, 0.6) | 0.1 (0.0, 0.3) |
|  | Zimbabwe | 2.7 (1.3, 5.7) | 0.4 (0.2, 0.9) | 0.2 (0.1, 0.5) | 0.1 (0.0, 0.3) |
|  | ***Pooled region*** | 3.4 (2.3, 5.1) | 0.8 (0.5, 1.2) | 0.4 (0.2, 0.8) | 0.2 (0.1, 0.4) |
| **WCA** | Benin | 8.4 (4.3, 18.9) | 3.9 (1.9, 7.6) | 0.7 (0.3, 1.5) | 0.6 (0.2, 1.7) |
|  | Burkina Faso | 5.4 (2.6, 10.6) | 3.4 (1.7, 6.4) | 0.7 (0.3, 1.7) | 0.5 (0.2, 1.3) |
|  | Cent. Afr. Rep. | 5.7 (2.8, 12.3) | 2.9 (1.4, 6.0) | 0.3 (0.1, 0.8) | 0.4 (0.1, 1.2) |
|  | Côte d'Ivoire | 5.3 (2.4, 14.0) | 2.3 (1.1, 4.8) | 0.3 (0.1, 0.8) | 0.3 (0.1, 1.1) |
|  | Cameroon | 14.4 (5.1, 42.3) | 2.9 (1.2, 7.0) | 0.6 (0.2, 1.9) | 0.3 (0.1, 1.3) |
|  | Dem. Rep. Congo | 2.4 (1.0, 5.3) | 2.4 (1.1, 5.2) | 0.2 (0.1, 0.6) | 0.3 (0.1, 1.3) |
|  | Congo | 1.6 (0.8, 3.8) | 1.6 (0.9, 3.0) | 0.3 (0.1, 0.8) | 0.2 (0.1, 0.5) |
|  | Gabon | 7.8 (4.1, 14.7) | 5.3 (2.3, 10.6) | 0.7 (0.3, 1.9) | 1.0 (0.3, 2.9) |
|  | Ghana | 1.8 (0.4, 7.9) | 1.4 (0.3, 5.1) | 0.1 (0.0, 0.7) | 0.1 (0.0, 0.9) |
|  | Guinea | 3.0 (0.9, 9.7) | 1.8 (0.6, 4.7) | 0.4 (0.1, 1.3) | 0.2 (0.0, 1.0) |
|  | Gambia | 3.8 (1.0, 13.7) | 2.7 (0.9, 8.2) | 0.3 (0.1, 1.1) | 0.4 (0.1, 1.8) |
|  | G. Bissau | 4.0 (0.9, 18.2) | 3.6 (1.2, 10.9) | 0.6 (0.1, 3.2) | 0.4 (0.1, 2.6) |
|  | Eq. Guinea | 1.0 (0.4, 2.2) | 5.5 (3.1, 10.2) | 0.4 (0.1, 0.9) | 0.6 (0.2, 1.6) |
|  | Liberia | 3.4 (1.7, 7.3) | 1.7 (0.8, 3.5) | 0.5 (0.2, 1.2) | 0.3 (0.1, 0.7) |
|  | Mali | 5.0 (1.7, 18.1) | 4.2 (1.8, 9.5) | 0.8 (0.3, 2.2) | 0.6 (0.2, 2.0) |
|  | Niger | 4.7 (2.3, 10.0) | 3.9 (2.1, 7.2) | 0.7 (0.2, 2.0) | 0.4 (0.1, 1.1) |
|  | Nigeria | 29.3 (11.9, 73.2) | 9.7 (4.0, 21.5) | 2.4 (0.7, 7.6) | 1.1 (0.3, 3.8) |
|  | Senegal | 8.5 (4.4, 15.8) | 3.9 (2.0, 7.2) | 0.5 (0.2, 1.3) | 0.5 (0.2, 1.5) |
|  | S. Leone | 3.8 (1.4, 10.9) | 1.3 (0.5, 3.9) | 0.8 (0.3, 2.4) | 0.3 (0.1, 1.0) |
|  | Chad | 10.3 (3.7, 27.0) | 19.1 (9.0, 37.0) | 2.3 (0.7, 6.3) | 2.6 (0.7, 7.9) |
|  | Togo | 4.0 (2.0, 8.4) | 3.5 (1.9, 6.7) | 0.5 (0.2, 1.3) | 0.4 (0.1, 1.2) |
|  | ***Pooled region*** | 6.6 (4.1, 10.5) | 3.9 (2.5, 5.9) | 0.5 (0.3, 1.0) | 0.5 (0.2, 1.2) |
| **SSA** | ***Regional Average*** | 4.0 (2.7, 5.7) | 1.4 (0.9, 2.0) | 0.5 (0.3, 0.8) | 0.2 (0.1, 0.5) |

FSW: female sex workers; MSM: men who have sex with men; PWID: people who inject drugs; TGW: transgender women; PLHIV: people living with HIV; ESA: Eastern and Southern Africa; WCA: Western and Central Africa; SSA: sub-Saharan Africa

### Supplementary Table S16: HIV prevalence and ART coverage sensitivity analysis to age group denominator

| **Region** | **Total population estimate (%)** | **MSM estimate (%; 95%CI)** | | **FSW estimate (%; 95%CI)** | |
| --- | --- | --- | --- | --- | --- |
|  |  | **15 to 49 years** | **15 to 29 years** | **15 to 49 years** | **15 to 39 years** |
| **HIV prevalence** | | | |  |  |
| ESA | 1% | 5 (3, 9) | 6 (3, 11) | 14 (10, 19) | 16 (12, 21) |
|  | 15% | 21 (12, 31) | 17 (9, 28) | 53 (45, 62) | 47 (38, 57) |
| WCA | 1% | 13 (9, 18) | 12 (9, 17) | 9 (7, 11) | 10 (7, 13) |
|  | 7% | 20 (13, 30) | 22 (14, 32) | 12 (9, 16) | 12 (9, 16) |
| **ART coverage** | | | |  |  |
| SSA | 40% | 44 (35, 54) | 45 (36, 52) | 32 (24, 41) | 35 (27, 44) |
| SSA | 80% | 61 (50, 70) | 61 (52, 68) | 69 (61, 76) | 68 (60, 74) |

HIV prevalence in men who have sex with men (MSM) and female sex workers (FSW) presented at 1% and 15% total population HIV prevalence in Eastern and Southern Africa (ESA) and at 1% and 7% total population HIV prevalence in Western and Central Africa (WCA). ART coverage in MSM presented at 40% and 80% total population ART coverage in sub-Saharan Africa (SSA). KP estimates using ages 15-49 as matched total population age group for HIV prevalence and ART coverage are as in primary analysis.

### Supplementary Table S17: GATHER checklist

| **Item**  **#** | **Checklist item** | **Reported on page #** |
| --- | --- | --- |
| **Objectives and funding** | | |
| **1** | Define the indicator(s), populations (including age, sex, and geographic entities), and time period(s) for which estimates were made. | 9-14 |
| **2** | List the funding sources for the work. | 15, 27 |
| **Data Inputs** | | |
| *For all data inputs from multiple sources that are synthesized as part of the study:* | | |
| **3** | Describe how the data were identified and how the data were accessed. | 9, 10. Supplementary Table S1 |
| **4** | Specify the inclusion and exclusion criteria. Identify all ad‐hoc exclusions. | 9, 10 |
| **5** | Provide information on all included data sources and their main characteristics. For each data source used, report reference information or contact name/institution, population represented, data collection method, year(s) of data collection, sex and age range, diagnostic criteria or measurement method, and sample size, as relevant. | 9, 10. Supplementary Table S1, S2 |
| **6** | Identify and describe any categories of input data that have potentially important biases (e.g., based on characteristics listed in item 5). | 10-12 |
| *For data inputs that contribute to the analysis but were not synthesized as part of the study:* | | |
| **7** | Describe and give sources for any other data inputs. | 9-14 |
| *For all data inputs:* | | |
| **8** | Provide all data inputs in a file format from which data can be efficiently extracted (e.g., a spreadsheet rather than a PDF), including all relevant meta‐data listed in item 5. For any data inputs that cannot be shared because of ethical or legal reasons, such as third‐party ownership, provide a contact name or the name of the institution that retains the right to the data. | Supplementary File 2 |
| **Data analysis** | | |
| **9** | Provide a conceptual overview of the data analysis method. A diagram may be helpful. | Figure 1, 9 |
| **10** | Provide a detailed description of all steps of the analysis, including mathematical formulae. This description should cover, as relevant, data cleaning, data pre‐processing, data adjustments and weighting of data sources, and mathematical or statistical model(s). | Figure 1, Supplementary Text S1 |
| **11** | Describe how candidate models were evaluated and how the final model(s) were selected. | N/A |
| **12** | Provide the results of an evaluation of model performance, if done, as well as the results of any relevant sensitivity analysis. | Supplementary Figures S2, S4, S12 |
| **13** | Describe methods for calculating uncertainty of the estimates. State which sources of uncertainty were, and were not, accounted for in the uncertainty analysis. | 14 |
| **14** | State how analytic or statistical source code used to generate estimates can be accessed. | 26 |
| **Results and Discussion** | | |
| **15** | Provide published estimates in a file format from which data can be efficiently extracted. | Supplementary Tables 9-11 |
| **16** | Report a quantitative measure of the uncertainty of the estimates (e.g. uncertainty intervals). | 15-19 |
| **17** | Interpret results in light of existing evidence. If updating a previous set of estimates, describe the reasons for changes in estimates. | 22 |
| **18** | Discuss limitations of the estimates. Include a discussion of any modelling assumptions or data limitations that affect interpretation of the estimates. | 23-24 |
